# Effectiveness of mRNA COVID-19 vaccine booster doses against Omicron severe outcomes

**DOI:** 10.1101/2022.10.31.22281766

**Authors:** Ramandip Grewal, Lena Nguyen, Sarah A Buchan, Sarah E Wilson, Sharifa Nasreen, Peter C. Austin, Kevin A. Brown, Deshayne B. Fell, Jonathan Gubbay, Kevin L. Schwartz, Mina Tadrous, Kumanan Wilson, Jeffrey C Kwong, Canadian Immunization Research Network (CIRN) Provincial Collaborative Network (PCN) Investigators

**Author notes:** **Correspondence to:** Jeffrey C Kwong, Senior Scientist, ICES, G1 06, 2075 Bayview Avenue, Toronto, Ontario, Canada, M4N 3M5, (or @DrJeffKwong on Twitter.

## Abstract

**Background:** To inform planning for further booster doses of COVID-19 vaccines, we estimated the effectiveness of monovalent mRNA vaccines against Omicron-associated severe outcomes in adults over time.

**Methods:** We used a test-negative design and multivariable logistic regression to estimate vaccine effectiveness (VE; 2, 3, or 4 doses compared to unvaccinated individuals) and marginal effectiveness (3 or 4 doses compared to 2 doses) against Omicron-associated hospitalization or death among community-dwelling adults aged ≥50 years who were tested for SARS-CoV-2 between January 2, 2022 and October 1, 2022 in Ontario, Canada, stratified by age group and time since vaccination. We also compared VE during periods of Omicron BA.1/BA.2 and BA.4/BA.5 sublineage predominance.

**Results:** We included 11,160 cases of Omicron-associated severe outcomes and 62,880 test-negative symptomatic controls. Depending on the age group, compared to unvaccinated individuals, VE was 91-98% 7-59 days after a third dose, waned to 76-87% after ≥240 days, was restored to 92-97% 7-59 days after a fourth dose, and waned to 86-89% after ≥120 days. Trends in marginal effectiveness were consistent with VE estimates. VE was lower during the BA.4/BA.5-predominant period compared to the BA.1/BA.2-predominant period based on the same intervals since vaccination.

**Conclusion:** Our findings suggest that 1 or 2 booster doses of monovalent mRNA COVID-19 vaccines initially restored very strong protection against Omicron-associated severe outcomes in all age groups, but VE subsequently declined over time with some age-related differences, and particularly so during a period of BA.4/BA.5 predominance.

## INTRODUCTION

COVID-19 vaccines first became available in Ontario, Canada in December 2020. Due to concerns about waning protection from the primary series and the emergence of more transmissible SARS-CoV-2 variants, third doses (first boosters) were offered to high-risk groups, including community-dwelling adults aged ≥70 years in November 2021.^1^ With the emergence of Omicron, the most transmissible and immune-evasive variant to date,^2^ third dose eligibility was expanded to all adults in December 2021.^3^ Ontario began offering fourth doses (second boosters) to adults aged ≥60 years in April 2022,^4^ and to all adults in July 2022.^5^ Booster dose policies differed for residents of long-term care facilities.^6^ Bivalent COVID-19 vaccines were introduced to Canadian vaccination programs starting in September 2022 and are now preferred,^7,8^ but monovalent vaccines are still authorized for use as boosters and are the products that have been most commonly received to date.

Various Omicron sublineages have circulated during 2022, with BA.1 and BA.2 predominating until June, and BA.4 and BA.5 predominating subsequently. The seroprevalence of prior SARS-CoV-2 infection increased substantially in Ontario during the Omicron period, from 6.4% in early January 2022 to 57.9% by end of August 2022.^9^

Due to increased transmissibility and immune evasion of emerging Omicron sublineages, more evidence is needed on the long-term effectiveness of booster doses of monovalent mRNA vaccines among older adults to inform planning for subsequent boosters and future shifts in vaccine development. Thus, we sought to estimate vaccine effectiveness (VE) of 2, 3, and 4 doses compared to unvaccinated individuals, and marginal effectiveness of 3 or 4 doses compared to 2 doses, in preventing severe outcomes (hospitalization or death) among community-dwelling adults aged ≥50 years during an Omicron-dominant period. We estimated marginal effectiveness due to concerns about differences between unvaccinated and vaccinated populations. We also sought to determine how VE varied during periods of BA.1/BA.2 versus BA.4/BA.5 predominance.

## METHODS

### Study design, setting, population, and data sources

Similar to past studies on COVID-19 VE in Ontario,^6,10,11^ we applied a test-negative design to provincial SARS-CoV-2 laboratory testing, COVID-19 vaccination, and health administrative datasets. These datasets were linked using unique encoded identifiers and analyzed at ICES (formerly the Institute of Clinical Evaluative Sciences).

We included community-dwelling adults aged ≥50 years who had ≥1 reverse-transcription polymerase chain reaction (RT-PCR) test for SARS-CoV-2 between January 2, 2022 and October 1, 2022. We excluded immunocompromised individuals (n=11,514) and those who received a bivalent mRNA vaccine (n=2,433), Ad26.COV2 (n=83), or >1 dose of ChAdOx1-S (n=1,043) by the index date (Figure S1). Approximately 95% of subjects received mRNA vaccines (mRNA-1273 or BNT162b2) for all doses. Omicron represented nearly 100% of all positive samples by late January 2022.^12–14^ Delta (B.1.617.2) cases identified using whole genome sequencing or based on an S-gene target positive screening result before January 24, 2022 (n=72) were excluded.

### Outcome and sampling strategy

The case definition was COVID-19-associated hospitalization or death due to, or partially due to, COVID-19, as specified by data entry guidelines for the public health COVID-19 surveillance database.^15^ We excluded hospitalizations when specimen collection occurred >3 days after admission and those flagged as being nosocomial. We sampled cases and controls by week of test, thus individuals could enter the study repeatedly, but once an individual became a case, they could not re-enter the study. Controls had to be symptomatic and test negative for SARS-CoV-2, but may or may not have had a severe outcome. We employed this sampling strategy to ensure the distribution of time of testing was consistent between cases and controls. The index date was the earliest of specimen collection, hospitalization, or death.

### COVID-19 vaccination

We classified community-dwelling adults by the number of doses received and time since most recent vaccination relative to the index date. For 2, 3, and 4 doses, we explored up to ≥300 days, ≥240 days, and ≥120 days post-vaccination, respectively. For booster doses of mRNA-1273, a half dose (50 mcg) was recommended for those younger than 70 years and a full dose (100 mcg) for those aged ≥70 years.^16^

### Statistical analysis

We used means and proportions to describe our sample by comparing: 1) test-negative symptomatic controls to test-positive Omicron cases who were hospitalized or died; 2) unvaccinated individuals to those who had received 2, 3, or 4 doses; and 3) individuals who had received 2 doses (≥7 days ago) to those who had received 3 or 4 doses. We used standardized differences (SD) to quantify the differences between groups.

Stratified by age group (50-59, 60-69, 70-79, ≥80 years), we used multivariable logistic regression to compare the odds of vaccination in cases to test-negative controls while adjusting for sex, age (continuous), public health unit region, four area-level variables representing different socio-demographic characteristics (household income quintile, essential worker quintile, persons per dwelling quintile, self-identified visible minority quintile), influenza vaccination during 2019-2020 or 2020-2021 (proxy for health behaviours), SARS-CoV-2 infection >90 days prior, number of SARS-CoV-2 tests within 3 months prior to December 14, 2020 (proxy for healthcare workers), comorbidities, receipt of home care services, and week of test. We estimated the logistic regression models using generalized estimating equations (GEE) with an exchangeable correlation structure since controls could be in a model more than once (13% of controls) leading to non-independence of observations. We calculated both VE and marginal effectiveness using the formula: (1-adjusted odds ratio)*100%.

To examine VE against various Omicron sublineages, we included in our multivariable models an interaction term for time period (BA.1/BA.2-predominant period: January 2, 2022 to July 2, 2022; BA.4/BA.5-predominant period: July 3, 2022 to October 1, 2022) (Figure 1).^14^ Among these sublineages, the distributions were approximately 50% BA.1 and 50% BA2 during the BA.1/BA.2-predominant period and 10% BA.4 and 90% BA.5 during the BA.4/BA.5-predominant period.^14^ GEE methods were not used in this analysis due to issues with convergence. For the estimates that did converge, GEE and non-GEE estimates and 95% confidence intervals were nearly identical.

**Figure 1:**
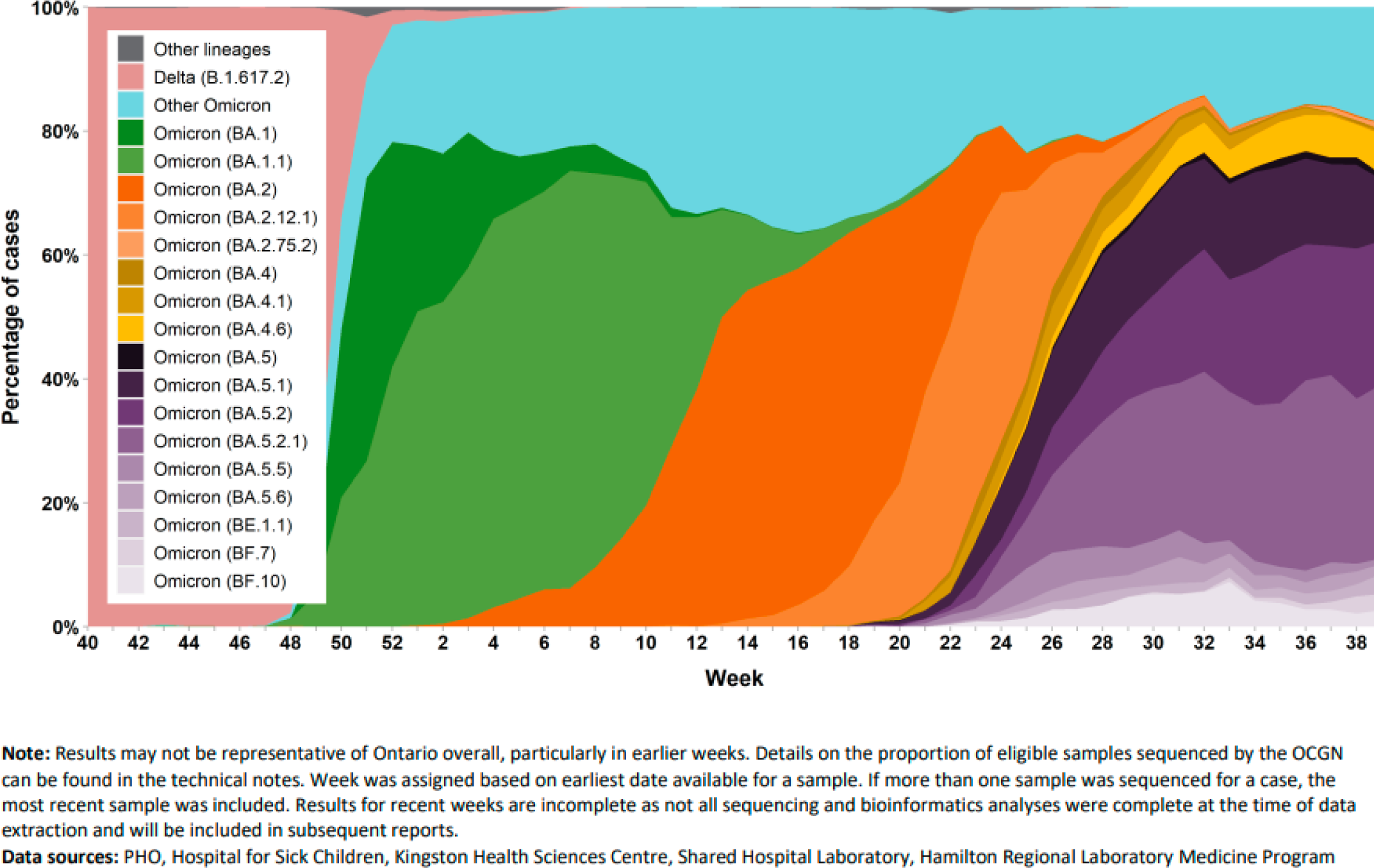
Percentage of COVID-19 cases by the most prevalent lineages and week, representative surveillance, Ontario, October 3, 2021 to October 1, 2022^14^

We used SAS version 9.1 (SAS Institute Inc., Cary, NC) for all analyses. All tests were 2-sided and we used p<0.05 as the level of significance. A SDs ≥0.1 were considered clinically relevant.

### Ethics approval

ICES is a prescribed entity under Ontario’s Personal Health Information Protection Act (PHIPA). Section 45 of PHIPA authorizes ICES to collect personal health information, without consent, for the purpose of analysis or compiling statistical information with respect to the management of, evaluation or monitoring of, the allocation of resources to or planning for all or part of the health system. Projects that use data collected by ICES under section 45 of PHIPA, and use no other data, are exempt from REB review. The use of the data in this project is authorized under section 45 and approved by ICES’ Privacy and Legal Office.

## RESULTS

We included 11,160 Omicron-associated severe outcomes and 62,880 symptomatic test-negative controls. Cases were older than controls (mean age of 76.7 years versus 65.7 years; standardized difference [SD]=0.93) (Table 1). More cases than controls were male (56.7% versus 37.4%; SD=0.40) and had at least one comorbid condition (92.7% versus 75.0%; SD=0.50). A higher proportion of cases versus controls were from areas with the lowest income quintile (26.8% versus 20.9%; SD=0.14) and were unvaccinated (28.0% versus 4.6%; SD=0.67). Compared to unvaccinated individuals, more vaccinated individuals were female, had previously received influenza vaccines, and fewer were from areas with the lowest income quintile (Table S2). More individuals with 2 versus 3 or 4 doses were from areas with the lowest income and fewer had received influenza vaccines (Table S3).

**Table 1:** Descriptive characteristics of community-dwelling adults aged ≥50 years tested for SARS-CoV-2 and with severe outcomes between January 2, 2022 and October 1, 2022 in Ontario, Canada, comparing Omicron-associated severe outcome cases to SARS-CoV-2 negative controls

|  | Omicron cases,<br>n (%) | SARS-CoV-2 negative<br>controls, n (%)<sup>a</sup> | SD<sup>b</sup> |
| --- | --- | --- | --- |
| **Total** | 11,160 | 62,880 |  |
| Characteristics |  |  |  |
| Age (years), mean (standard deviation) | 76.65 ± 11.69 | 65.67 ± 11.97 | 0.93 |
| 50-59 | 1,136 (10.2%) | 25,002 (39.8%) | 0.73 |
| 60-69 | 1,926 (17.3%) | 16,139 (25.7%) | 0.21 |
| 70-79 | 2,975 (26.7%) | 11,585 (18.4%) | 0.20 |
| $\geq 80$ | 5,123 (45.9%) | 10,154 (16.1%) | 0.68 |
| Male sex | 6,331 (56.7%) | 23,494 (37.4%) | 0.40 |
| Public health unit region |  |  |  |
| Central East | 691 (6.2%) | 4,494 (7.1%) | 0.04 |
| Central West | 2,184 (19.6%) | 9,626 (15.3%) | 0.11 |
| Durham | 309 (2.8%) | 2,889 (4.6%) | 0.10 |
| Eastern | 932 (8.4%) | 2,634 (4.2%) | 0.17 |
| North | 936 (8.4%) | 12,837 (20.4%) | 0.35 |
| Ottawa | 527 (4.7%) | 1,226 (1.9%) | 0.15 |
| Peel | 1,275 (11.4%) | 7,035 (11.2%) | 0.01 |
| South West | 1,449 (13.0%) | 11,654 (18.5%) | 0.15 |
| Toronto | 1,969 (17.6%) | 7,628 (12.1%) | 0.16 |
| York | 845 (7.6%) | 2,640 (4.2%) | 0.14 |
| Missing | 43 (0.4%) | 217 (0.3%) | 0.01 |
| Household income quintile |  |  |  |
| 1 (lowest) | 2,986 (26.8%) | 13,139 (20.9%) | 0.14 |
| 2 | 2,480 (22.2%) | 12,878 (20.5%) | 0.04 |
| 3 | 2,128 (19.1%) | 12,134 (19.3%) | 0.01 |
| 4 | 1,930 (17.3%) | 12,115 (19.3%) | 0.05 |
| 5 (highest) | 1,598 (14.3%) | 12,414 (19.7%) | 0.14 |
| Missing | 38 (0.3%) | 200 (0.3%) | 0.00 |
| Essential workers quintile |  |  |  |
| 1 (0%–32.5%) | 1,566 (14.0%) | 9,688 (15.4%) | 0.04 |
| 2 (32.5%–42.3%) | 2,263 (20.3%) | 13,621 (21.7%) | 0.03 |
| 3 (42.3%–49.8%) | 2,364 (21.2%) | 13,731 (21.8%) | 0.02 |
| 4 (50.0%–57.5%) | 2,438 (21.8%) | 12,964 (20.6%) | 0.03 |
| 5 (57.5%–100%) | 2,457 (22.0%) | 12,397 (19.7%) | 0.06 |
| Missing | 72 (0.6%) | 479 (0.8%) | 0.01 |
| Persons per dwelling quintile |  |  |  |
| 1 (0–2.1) | 2,919 (26.2%) | 14,564 (23.2%) | 0.07 |
| 2 (2.2–2.4) | 2,166 (19.4%) | 13,979 (22.2%) | 0.07 |
| 3 (2.5–2.6) | 1,592 (14.3%) | 8,512 (13.5%) | 0.02 |
| 4 (2.7–3.0) | 2,229 (20.0%) | 13,146 (20.9%) | 0.02 |
| 5 (3.1–5.7) | 2,172 (19.5%) | 12,174 (19.4%) | 0.00 |
| Missing | 82 (0.7%) | 505 (0.8%) | 0.01 |
| Self-identified visible minority quintile |  |  |  |
| 1 (0.0%–2.2%) | 2,109 (18.9%) | 14,855 (23.6%) | 0.12 |
| 2 (2.2%–7.5%) | 1,976 (17.7%) | 14,049 (22.3%) | 0.12 |
| 3 (7.5%–18.7%) | 2,017 (18.1%) | 11,347 (18.0%) | 0.00 |
| 4 (18.7%–43.5%) | 2,202 (19.7%) | 10,519 (16.7%) | 0.08 |
| 5 (43.5%–100%) | 2,785 (25.0%) | 11,631 (18.5%) | 0.16 |
| Missing | 71 (0.6%) | 479 (0.8%) | 0.02 |
| Receipt of 2019-2020 and/or<br>2020-2021 influenza vaccination | 5,931 (53.1%) | 32,573 (51.8%) | 0.03 |
| Prior positive SARS-CoV-2 test | 128 (1.1%) | 3,056 (4.9%) | 0.22 |
| Number of SARS-CoV-2 tests within 3<br>months prior to December 14, 2020 |  |  |  |
| 0 | 9,875 (88.5%) | 45,885 (73.0%) | 0.40 |
| 1 | 890 (8.0%) | 8,995 (14.3%) | 0.20 |
| ≥2 | 395 (3.5%) | 8,000 (12.7%) | 0.34 |
| Any comorbidity | 10,344 (92.7%) | 47,133 (75.0%) | 0.50 |
| Receipt of home care services |  |  |  |
| None | 10,112 (90.6%) | 59,641 (94.8%) | 0.16 |
| Short stay | 371 (3.3%) | 1,652 (2.6%) | 0.04 |
| Long stay | 630 (5.6%) | 1,441 (2.3%) | 0.17 |
| Palliative | 47 (0.4%) | 146 (0.2%) | 0.03 |
| Unvaccinated<sup>c</sup> | 3,127 (28.0%) | 2,894 (4.6%) | 0.67 |
| Time since second dose<sup>c</sup> |  |  |  |
| 7-59 days | 36 (0.3%) | 231 (0.4%) | 0.01 |
| 60-119 days | 125 (1.1%) | 659 (1.0%) | 0.01 |
| 120-179 days | 409 (3.7%) | 1,950 (3.1%) | 0.03 |
| 180-239 days | 1,216 (10.9%) | 4,437 (7.1%) | 0.13 |
| 240-299 days | 476 (4.3%) | 2,019 (3.2%) | 0.06 |
| ≥300 days | 751 (6.7%) | 2,911 (4.6%) | 0.09 |
| Time since third dose<sup>c</sup> |  |  |  |
| 0-6 days | 101 (0.9%) | 809 (1.3%) | 0.04 |
| 7-59 days | 643 (5.8%) | 12,928 (20.6%) | 0.45 |
| 60-119 days | 971 (8.7%) | 11,618 (18.5%) | 0.29 |
| 120-179 days | 922 (8.3%) | 8,786 (14.0%) | 0.18 |
| 180-239 days | 875 (7.8%) | 4,703 (7.5%) | 0.01 |
| ≥240 days | 447 (4.0%) | 2,186 (3.5%) | 0.03 |
| Time since fourth dose<sup>c</sup> |  |  |  |
| 0-6 days | 43 (0.4%) | 313 (0.5%) | 0.02 |
| 7-59 days | 269 (2.4%) | 2,971 (4.7%) | 0.13 |
| 60-119 days | 404 (3.6%) | 2,302 (3.7%) | 0.00 |
| ≥120 days | 345 (3.1%) | 1,163 (1.8%) | 0.08 |

\*Note, not unique by person; individuals may be included more than once.
<sup>a</sup>Proportion reported, unless stated otherwise.
<sup>b</sup>SD=standardized difference. Standardized differences of >0.10 are considered clinically relevant. Comparing Omicron cases to test-negative controls.
<sup>c</sup>Sum of all rows (unvaccinated and vaccinated) equals 100%.

Compared to unvaccinated individuals, VE against severe disease increased shortly after receipt of booster doses but subsequently declined over time (Figure 2, Tables S4-S5). For example, among subjects aged 70-79 years, VE decreased from: 84% (95%CI, 57-94%) 7-59 days after a second dose to 71% (95%CI, 63-78%) after ≥300 days; 96% (95%CI, 95-97%) 7-59 days after a third dose to 79% (95%CI, 71-85%) after ≥240 days; and 93% (95%CI, 91-95%) 7-59 days after a fourth dose to 89% (95%CI, 84-92%) after ≥120 days. The decline in VE after a third dose appeared to plateau after 180 days. VE was generally lower with increasing age.

**Figure 2:**
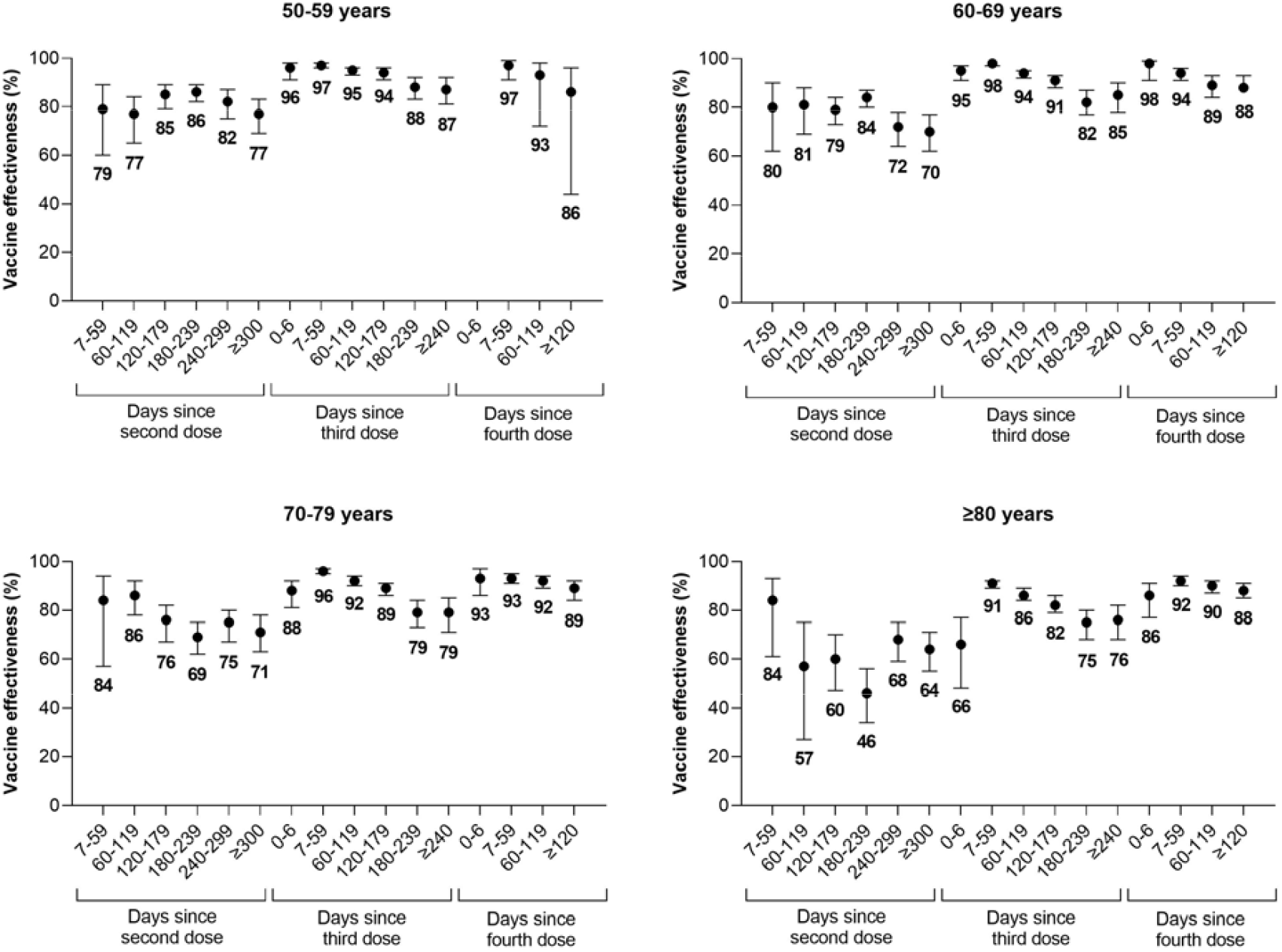
Vaccine effectiveness of 2, 3, and 4 doses of monovalent mRNA COVID-19 vaccines against Omicron-associated severe outcomes among community-dwelling adults aged ≥50 years in Ontario, Canada, compared to unvaccinated adults (Note: Estimates were not reported if they were unstable [i.e., 95% confidence interval width exceeded 100 percentage points]).

Marginal effectiveness peaked 7-59 days after third and fourth doses and declined over time. Once again, among adults aged 70-79 years, compared to 2 doses (median 218 days since a second dose), the marginal effectiveness of a third dose decreased from 83% (95%CI, 79-86%) 7-59 days after a third dose to 45% (95%CI, 24-59%) after ≥240 days and the marginal effectiveness of a fourth dose decreased from 80% (95%CI, 74-85%) 7-59 days after a fourth dose to 69% (95%CI, 57-78%) after ≥120 days (Figure 3, Table S6).

**Figure 3:**
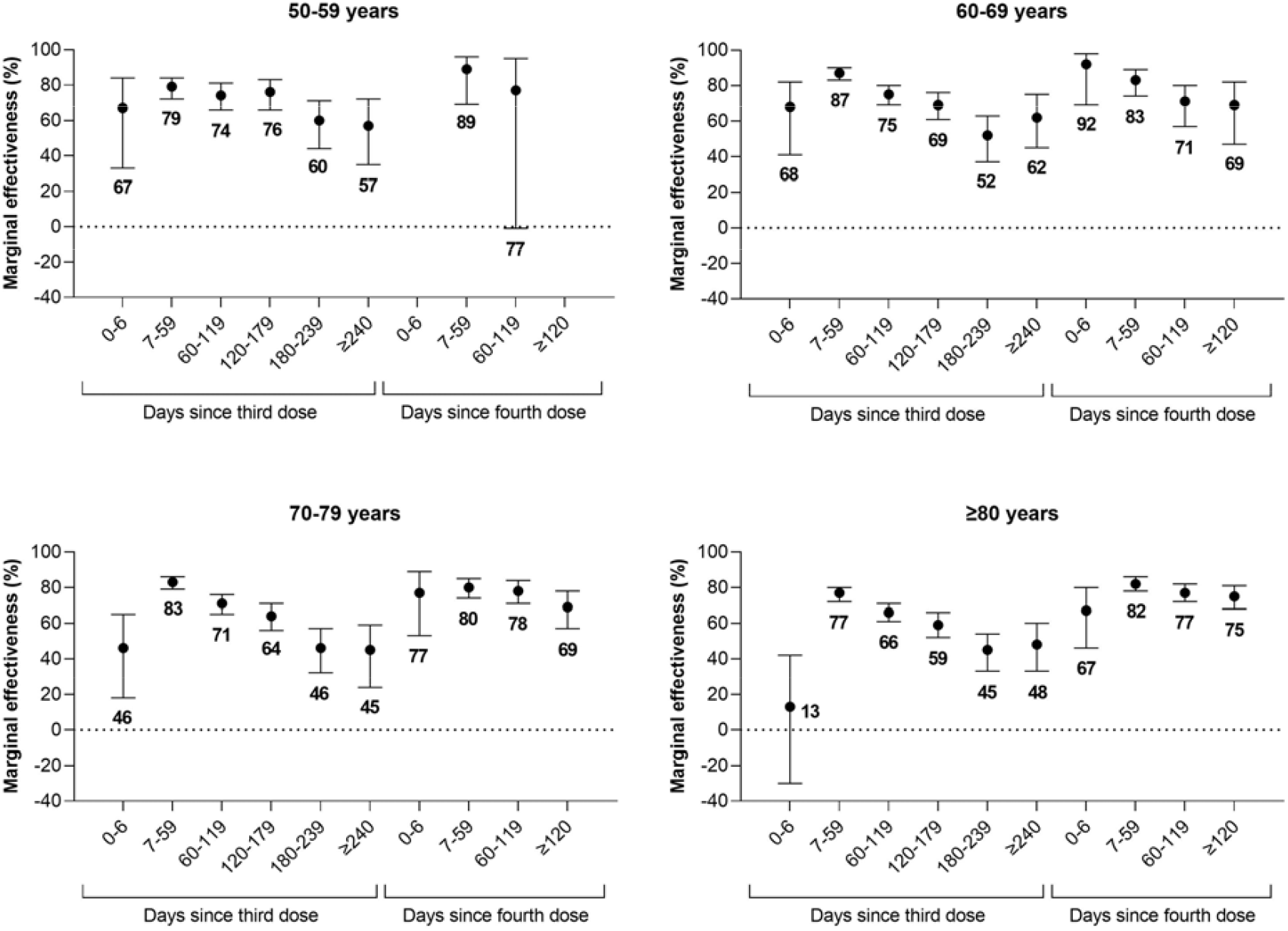
Marginal effectiveness of a third and fourth dose of monovalent mRNA COVID-19 vaccines against Omicron-associated severe outcomes among community-dwelling adults aged ≥50 years in Ontario, Canada, compared to adults who received 2 doses (Note: Estimates were not reported if they were unstable [i.e., 95% confidence interval width exceeded 100 percentage points]).

VE estimates were lower during the BA.4/BA.5-predominant period compared to the BA.1/BA.2-predominant period, with differences widening as time since vaccination increased (Figure 4, Table S7). For example, among subjects aged 70-79 years, VE 7-59 days after a third dose was 96% (95%CI, 96-97%) (median 34 days since a third dose) during the BA.1/BA.2-predominant period compared to 86% (95%CI, 37-97%) (median 41 days since a third dose) during the BA.4/BA.5-predominant period (p=0.08 for the between-period interaction), whereas VE 180-239 days after a third dose was 91% (95%CI, 85-95%) (median 189 days since a third dose) during the BA.1/BA.2-predominant period compared to 59% (95%CI, 44-70%) (median 214 days since a third dose) during the BA.4/BA.5-predominant period (p<0.001 for the between-period interaction).

**Figure 4:**
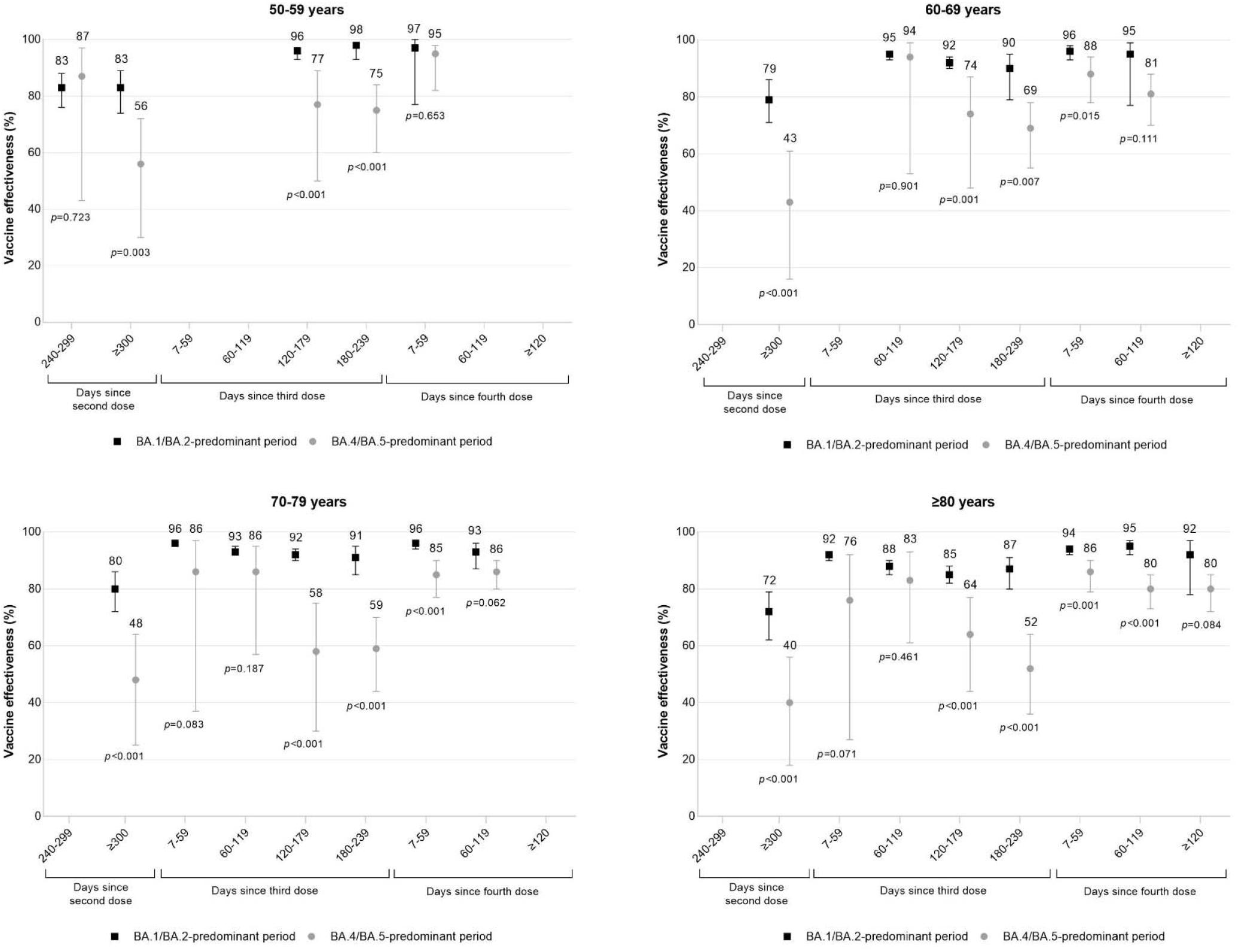
Vaccine effectiveness against Omicron-associated severe outcomes among community-dwelling adults aged ≥50 years in Ontario, Canada, comparing those who received ≥2 doses of monovalent mRNA COVID-19 vaccines to those who received none, by age and time since vaccination, during periods of BA.1/BA.2 (January 2 to July 2, 2022) and BA.4/BA.5 (July 3 to October 1, 2022) predominance (Note: Estimates were not reported if they were unstable for either period).

## DISCUSSION

Among community-dwelling adults aged ≥50 years in Ontario, VE against Omicron-associated severe outcomes increased with booster doses of monovalent mRNA COVID-19 vaccines, but protection waned over time after each dose. Third doses continued to provide strong protection (85-87%) against severe outcomes among individuals aged 50-69 years even 8 months after vaccination, but lower protection (76-79%) among adults aged ≥70 years. Fourth doses restored waning of protection from third doses and continued to provide strong protection (86-89%) 4 months after vaccination for all age groups. However, VE in the BA.4/BA.5-predominant period was lower than during the BA.1/BA.2-predominant period across the same time intervals after vaccination, especially with increasing time since vaccination.

Comparisons with other jurisdictions are challenging due to heterogeneity in study design, population characteristics, outcomes and exposures, vaccines, and observation periods. Our fourth dose VE estimates were slightly higher than those observed in the United States, where fourth dose VE against hospitalizations was 80% (95%CI, 71-85%) after ≥7 days among adults aged ≥50 years.^17^ Studies from Israel found that waning of protection against severe outcomes was significantly slower than against infection and that marginal effectiveness of booster doses against infection waned faster after fourth doses compared to third doses.^18–20^ They were unable to determine if trends were similar for severe outcomes due to the short follow-up period. In our study, the waning of protection against severe outcomes observed ≥120 days after a fourth dose was comparable to that seen 120-179 days after a third dose. Although differences in timing of vaccination within those time periods may influence VE estimates, the median time since vaccination was 141-145 days (depending on the age group) for the 120-179 days post third dose group and 140-147 days for the ≥120 days post fourth dose group, suggesting that waning of protection after a fourth dose may follow a similar trajectory as after a third dose.

Available evidence on VE of monovalent COVID-19 vaccines against severe outcomes caused by the BA.4/BA.5 Omicron sublineages varies. In the UK, compared to a second dose, marginal effectiveness of a third or fourth dose against BA.4/BA.5-versus BA.2-related hospitalizations was similar using the same time intervals since vaccination.^21^ Similarly, VE of a third dose against hospitalizations was comparable between BA.1/BA.2-predominant and BA.4/BA.5-predominant periods in South Africa.^22^ Conversely, in Portugal, 3-dose protection against severe outcomes was lower among BA.5 versus BA.2 cases.^23^ A study among Kaiser Permanente members found that VE of third and fourth doses against BA.4/BA.5-related hospitalizations was lower compared to BA.1/BA.2-related hospitalizations, whereas another study among individuals admitted to IVY Network hospitals saw this difference for a third dose but not for 2 or 4 doses.^24,25^ The IVY Network study reported 3-dose VE of 79% (95%CI, 74-84%) and 60% (95%CI, 12-81%), respectively, during the BA.1/BA.2-predominant versus BA.4/BA.5-predominant periods 7-120 days after vaccination.^25^

Potential explanations for lower VE during the period of BA.4/BA.5-predominance compared to BA.1/BA.2-predominance include longer intervals between booster dose receipt and outcomes, increased incidence of undocumented infections, and increased BA.4/BA.5 immune evasion.^17^ In our study, differences in the median numbers of days since booster receipt between the BA.1/BA.2-predominant versus BA.4/BA.5-predominant periods were always <30 days, so longer post-booster follow-up during the BA.4/BA.5-predominant period was unlikely to be a major contributor to the large observed differences in VE. VE may be underestimated in the setting of undocumented infections if unvaccinated individuals are more likely to be infected than vaccinated individuals because the former will have infection-induced immunity, and the extent of VE underestimation may increase as prior infections become more prevalent in the population. However, we noted that VE declined considerably faster as time since vaccination increased during the relatively brief BA.4/BA.5-predominant period (only 3 months) compared to the BA.1/BA.2-predominant period, suggesting that bias from undocumented prior infections is unlikely to account entirely for the differences. Therefore, among these potential explanations, increased immune evasion by BA.4/BA.5 sublineages is likely the largest contributor to these differences in VE.

Based on the evidence to date, the level of protection offered by bivalent vaccines remains unclear. Findings from a phase 2-3 trial suggest that Moderna’s bivalent vaccine elicits higher titres of neutralizing antibodies against Omicron sublineages BA.1 and BA.4/BA.5 compared to Moderna’s ancestral monovalent vaccine.^26^ In contrast, two observational immunogenicity studies found the Pfizer and Moderna bivalent vaccines elicited similar levels of neutralizing antibodies against BA.4/BA.5 as the ancestral monovalent vaccines.^27,28^

This study had some limitations. First, data on rapid antigen tests were not available, and this was the main source of testing after December 31, 2021, when eligibility for RT-PCR testing was restricted in Ontario to individuals considered at high risk of acquiring SARS-CoV-2.^29^ Thus, while we adjusted for prior SARS-CoV-2 infections documented by RT-PCR, we could not account for prior infections confirmed only by rapid antigen tests. This could bias VE estimates downward or upward depending on whether unvaccinated or vaccinated individuals are more likely to have prior undocumented infections. Second, because whole genome sequencing was not performed on all cases, we were unable to estimate VE against BA.1, BA.2, BA.4, and BA.5 separately but instead combined BA.1 and BA.2 during one period and BA.4 and BA.5 during another period based on when each grouping circulated. Last, there remains the potential for residual confounding since we were limited to the covariates available in the databases used. Unmeasured differences between unvaccinated and vaccinated individuals may introduce bias, but the consistency between the VE and marginal effectiveness estimates is reassuring. A significant strength of our study is the length of the follow-up period, allowing us to estimate VE ≥4 months after fourth doses. Also, unlike most other studies, we stratified our analyses by age group, which provides more refined VE estimates for decision-making.

Our findings suggest that while booster doses of monovalent mRNA COVID-19 vaccines initially restore strong protection against Omicron-associated hospitalizations and death among community-dwelling older adults and then subsequently wane over time, much uncertainty remains. Although protection remained strong 4 months after a fourth dose for all age groups, whether waning increases past this period remains unknown, and combined with the evidence of reduced VE against BA.4/BA.5 sublineages and the possibility that vaccines could be even less effective against newly emerging sublineages such as BQ.1.1 and XBB, subsequent boosters and other measures (e.g., face masks, improved ventilation, filtration of indoor air) may be needed to mitigate the impact of Omicron and future SARS-CoV-2 variants. It will be important to continue monitoring VE given the scarcity of VE data against BA.4/BA.5, newly emerging sublineages, and the introduction of bivalent vaccines.

## Data Availability

Data sharing agreement
The dataset from this study is held securely in coded form at ICES. While legal data sharing agreements between ICES and data providers (e.g., healthcare organizations and government) prohibit ICES from making the dataset publicly available, access may be granted to those who meet prespecified criteria for confidential access, available at www.ices.on.ca/DAS. The full dataset creation plan and underlying analytic code are available from the authors upon request, understanding that the computer programs may rely upon coding templates or macros that are unique to ICES and are therefore either inaccessible or may require modification.

## Acknowledgements

We would like to acknowledge Public Health Ontario for access to vaccination data from COVaxON, case-level data from CCM and COVID-19 laboratory data, as well as assistance with data interpretation. We also thank the staff of Ontario’s public health units who are responsible for COVID-19 case and contact management and data collection within CCM. We thank IQVIA Solutions Canada Inc. for use of their Drug Information Database. The authors are grateful to the Ontario residents without whom this research would be impossible.

## Funding and disclaimers

This work was supported by funding from the Canadian Immunization Research Network (CIRN) through a grant from the Public Health Agency of Canada and the Canadian Institutes of Health Research (CNF 151944), and also by funding from the Public Health Agency of Canada, through the Vaccine Surveillance Working Party and the COVID-19 Immunity Task Force. This study was supported by Public Health Ontario and by ICES, which is funded by an annual grant from the Ontario Ministry of Health (MOH) and Ministry of Long-Term Care (MLTC). This work was also supported by the Ontario Health Data Platform (OHDP), a Province of Ontario initiative to support Ontario’s ongoing response to COVID-19 and its related impacts. Jeffrey C. Kwong is supported by a Clinician-Scientist Award from the University of Toronto Department of Family and Community Medicine.

The study sponsors did not participate in the design and conduct of the study; collection, management, analysis and interpretation of the data; preparation, review or approval of the manuscript; or the decision to submit the manuscript for publication. Parts of this material are based on data and/or information compiled and provided by the Canadian Institute for Health Information (CIHI), Ontario Health, and by Cancer Care Ontario (CCO). However, the analyses, conclusions, opinions and statements expressed herein are solely those of the authors, and do not reflect those of the funding or data sources; no endorsement by ICES, MOH, MLTC, OHDP, its partners, the Province of Ontario, Ontario Health, CIHI, or CCO is intended or should be inferred.

## Data sharing agreement

The dataset from this study is held securely in coded form at ICES. While legal data sharing agreements between ICES and data providers (e.g., healthcare organizations and government) prohibit ICES from making the dataset publicly available, access may be granted to those who meet prespecified criteria for confidential access, available at www.ices.on.ca/DAS. The full dataset creation plan and underlying analytic code are available from the authors upon request, understanding that the computer programs may rely upon coding templates or macros that are unique to ICES and are therefore either inaccessible or may require modification.

