## Supplementary material for "Effectiveness of mRNA COVID-19 vaccine booster doses against Omicron severe outcomes"

**Supplementary Appendix**

This appendix has been provided by the authors to give readers additional information about their work.

Supplementary Text: Determination of symptom status at the time of SARS-CoV-2 testing

The Ontario Laboratories Information System (OLIS) contains open fields (specifically, using the Patient Note Clinical Information field or reporting under the observation code XON13543-4 [Patient symptoms]) that records whether individuals tested for SARS-CoV-2 presented with symptoms at the time of the test. These character-based fields were originally delimited by commas, slashes, semicolons, or ampersands. Character text-strings were parsed and aggregated.

One of the authors, JCK, identified symptom classifications available in OLIS as of October 5, 2022 that were likely to be due to COVID-19. Low-frequency terms (appearing <25 times throughout) were excluded. Values listed in the symptoms fields were classified as “symptomatic” and “asymptomatic.” We purposely chose a broadly inclusive definition to capture all potentially relevant COVID-19 symptoms, including atypical symptoms and chronic conditions, based on our scientific and medical understanding of COVID-19-related symptoms

Terms determined to be indicative of COVID-19 symptoms (classified as ‘symptomatic’) are listed below. In addition to this list, we used SYMPTOMATIC (or partial spellings thereof) or mention of symptom onset.

", *, +VE RAPID, +VE RAPID TEST, -, ., .ASY, 0, 0 TASTE, 0.97, 0.98, 0.99, 01, 02, 03, 04, 05, 06, 07, 08, 09, 1, 10, 100, 100%, 101, 102, 11, 12, 13, 14, 15, 16, 17, 18, 19, 2, 20, 2020, 2020	OUT PT, 2020 OUT PT, 2020...OUT PT, 2021, 2021	OUT PT, 2021	IN PT, 2021	OUT PT, 2021 IN PT, 2021 OUT PT, 2021 - COUGH, 2021 - FEVER, 2021 - SORE THROAT, 2021-11-05, 2021-11-16, 2021-11-17, 2021-11-18, 2021-12-21, 2021-12-22, 2021-12-25, 2021-12-26, 2021-12-27, 2021-12-28, 2021-12-29, 2021., 2022, 2022	IN PT, 2022	OUT PT, 2022 IN PT, 2022 OUT PT, 21, 21 - COUGH, 21., 22, 23, 24, 25, 26, 27, 28, 29, 2ND SWAB, 3, 30, 31, 35, 35.0, 35.1, 35.2, 35.3, 35.4, 35.5, 35.6, 35.7, 35.8, 35.9, 36, 36.0, 36.1, 36.2, 36.3, 36.4, 36.5, 36.6, 36.7, 36.8, 36.9, 37, 37.0, 37.1, 37.2, 37.3, 37.4, 37.5, 37.6, 37.7, 37.8, 37.9, 38, 38.0, 38.1, 38.2, 38.3, 38.4, 38.5, 38.6, 38.7, 38.8, 38.9, 39, 39.0, 39.1, 39.2, 39.3, 39.4, 39.5, 39.6, 3RD SWAB, 4, 40, 5, 6, 7, 8, 9, 97%, 98%, 99, 99%, A, A COLD, AB PAIN, ABD, ABD DISCOMFORT, ABD PAIN, ABD. PAIN, ABD.PAIN, ABDO CRAMPS, ABDO DISCOMFORT, ABDO PAIN, ABDO PAIN AND HEADACHE, ABDOMEN PAIN, ABDOMINAL, ABDOMINAL CRAMPING, ABDOMINAL CRAMPS, ABDOMINAL DISCOMFORT, ABDOMINAL PAIN, ABDOMINAL PAINS, ABDOMINAL UPSET, ABDOPAIN, ABNORMAL ULTRASOUND, AC VISITOR, ACHE, ACHES, ACHES AND CHILLS, ACHES AND PAIN, ACHES AND PAINS, ACHES CHILLS, ACHES FATIGUE, ACHES HEADACHE, ACHES RUNNY NOSE, ACHEY, ACHINESS, ACHING, ACHY, ACHY BODY, ACHY HEADACHE, ACHY JOINTS, ACHY MUSCLES, ACUTE STROKE, ADMISSION, ADMISSION ONLY, ADMISSION PROTOCOL, ADMISSION SWAB, ADMISSION TO HOSPICE, ADMISSION TO HOSPITAL, ADMISSON, ADMIT, ADMITTED, AFEBRILE, AFIB, ALLERGIES, ALLERGY SYMPTOMS, ALTERED LOC, ALTERED TASTE, AND HEADACHE, ANEMIA, ANOREXIA, ANOSMIA, ANXIETY, AP, APPENDICITIS, APPETITE, APRIL 22, APRIL 28, ARTHRALGIA, ARTHRITIS, AS, ASTHMA, ASX, ASY, ASYMPOTMATIC, ASYMPT., ASYMPTOMATIC, ASYMPTOMATIC - CLEARANCE, ASYMPTOMATIC - EXPOSURE, ASYMPTOMATIC - SURVEILLANCE, ASYPMTOMATIC, ASYPTOMATIC, AUG 30, AUTOPSY, AX CENTRE, AYMPTOMATIC, Anemia, Apr-22, Apr-28, B, BACK ACHE, BACK ACHES, BACK PAIN, BACKACHE, BACKPAIN, BAD HEADACHE, BARKING COUGH, BEING ADMITTED, BLEEDING, BLEEDING IN PREGNANCY, BLOATING, BLOODY STOOLS, BODY, BODY ACH, BODY ACHE, BODY ACHE AND HEADACHE, BODY ACHE HEADACHE, BODY ACHE., BODY ACHEA, BODY ACHES, BODY ACHES 12, BODY ACHES AND CHILLS, BODY ACHES AND HEADACHE, BODY ACHES AND PAINS, BODY ACHES CHILLS, BODY ACHES CONGESTION, BODY ACHES FATIGUE, BODY ACHES HEADACHE, BODY ACHES HEADACHES, BODY ACHES RUNNY NOSE, BODY ACHES., BODY ACHES. 2022, BODY CHILLS, BODY MALAISE, BODY PAIN, BODY PAINS, BODY RASH, BODY WEAKNESS, BODYACH, BODYACHE, BODYACHES, BODYACHES CHILLS, BODYPAIN, BOSY ACHES, BRAIN FOG, BREATHING DIFFICULTY, BREATHING ISSUES, BRONCHITIS, BURNING CHEST, BURNING EYES, BURNING IN CHEST, Bleeding in pregnancy, Bloating, Bloody stools, C, C-CASE, CAMP, CANCER, CARDIAC, CARDIAC ARREST, CAREGIVER, CATARACT, CCU), CENTRAL CAREGIVER FOR LONG-TERM CARE, CH, CHANGE IN TASTE, CHANGE IN TASTE AND SMELL, CHEST, CHEST AND NASAL CONGESTION, CHEST BURNING, CHEST COLD, CHEST CONG, CHEST CONGESTED, CHEST CONGESTION, CHEST DISCOMFORT, CHEST HEAVINESS, CHEST HEAVY, CHEST HURTS, CHEST INFECTION, CHEST IRRITATION, CHEST PAIN, CHEST PAINS, CHEST PRESSURE, CHEST SORE, CHEST SORENESS, CHEST TIGHNESS, CHEST TIGHT, CHEST TIGHTNESS, CHESTPAIN, CHF, CHI, CHIILS, CHIL, CHILL, CHILLA, CHILLLS, CHILLS, CHILLS ACHES, CHILLS AND BODY ACHE, CHILLS AND BODY ACHES, CHILLS AND FATIGUE, CHILLS AND HEADACHE, CHILLS AND RUNNY NOSE, CHILLS AND SWEATS, CHILLS BODY ACHE, CHILLS BODY ACHES, CHILLS CONGESTION, CHILLS DIARRHEA, CHILLS FATIGUE, CHILLS HEADACHE, CHILLS NAUSEA, CHILLS RUNNY NOSE, CHILLS SWEATS, CHILLS., CHILLS. 2021, CHILLS. 2022, CHILLS. HEADACHE, CHILS, CHRONIC COUGH, CLAMMY, CLEARANCE, CLEARING THROAT, CLOSE CONTACT, COGESTION, COGUH, COLD, COLD CHILLS, COLD FLASHES, COLD LIKE, COLD LIKE SYMPTOMS, COLD RUNNY NOSE, COLD SORE, COLD SWEAT, COLD SWEATS, COLD SX, COLD SYMPTOMS, COLD-LIKE SYMPTOMS, COLDS, CONFUSED, CONFUSION, CONG, CONGE, CONGEATION, CONGES, CONGESITON, CONGEST, CONGESTED, CONGESTED CHEST, CONGESTED COUGH, CONGESTED FATIGUE, CONGESTED HEADACHE, CONGESTED NOSE, CONGESTED RUNNY NOSE, CONGESTED SNEEZING, CONGESTED., CONGESTI, CONGESTIO, CONGESTION, CONGESTION 2021, CONGESTION ACHES, CONGESTION AND FATIGUE, CONGESTION AND HEADACHE, CONGESTION AND RUNNY NOSE, CONGESTION BODY ACHES, CONGESTION CHILLS, CONGESTION DIARRHEA, CONGESTION FATIGUE, CONGESTION FATIGUE HEADACHE, CONGESTION HEADACHE, CONGESTION HEADACHES, CONGESTION NAUSEA, CONGESTION RUNNY NOSE, CONGESTION RUNNY NOSE HEADACHE, CONGESTION SNEEZING, CONGESTION SOB, CONGESTION., CONGESTION. 2021, CONGESTION. 2022, CONGESTION. HEADACHE, CONGESTION. RUNNY NOSE, CONGESTIONS, CONGETION, CONGSTION, CONJESTION, CONJUCTION, CONJUCTIVITIS, CONJUNCTION, CONJUNCTIVITIS, CONSTIPATION, CONTACT, COPD, CORE THROAT, COUGH, COUGH 12, COUGH 2021, COUGH CONGESTION, COUGH DRY, COUGH FEVER, COUGH ONSET 20, COUGH PRODUCTIVE, COUGH RUNNY NOSE, COUGH SOB, COUGH SORE THROAT, COUGH SORE THROAT RUNNY NOSE, COUGH., COUGH. 2021, COUGH. 2022, COUGHING, COVID POSITIVE, COVID RESWAB, COVID TESTING, CP, CRAMPING, CRAMPS, CROUP, D, DAIRRHEA, DATE :2021, DATE OF SYMPTOM ONSET: 2021, DATE: 2021, DATE:2021, DD, DEC, DEC APPETITE, DEC.19, DEC.20, DEC.21, DEC.22, DEC.23, DEC.24, DEC.25, DEC.26, DEC.27, DEC.APPETITE, DECEASED, DECREASE APPETITE, DECREASE APPETITE. 2021, DECREASE TASTE, DECREASED APPETITE, DECREASED APPETITE., DECREASED LOC, DECREASED SMELL, DECREASED TASTE, DEHYDRATION, DELERIUM, DELIRIUM, DEMENTIA, DEPRESSION, DIA, DIAHERRA, DIAHHREA, DIAHREA, DIAHRREA, DIAPHORESIS, DIAPHORETIC, DIAR, DIAREHA, DIARHEA, DIARHHEA, DIARR, DIARREA, DIARREAH, DIARREHA, DIARREHEA, DIARRH, DIARRHE, DIARRHEA, DIARRHEA AND FATIGUE, DIARRHEA AND HEADACHE, DIARRHEA AND NAUSEA, DIARRHEA AND VOMITING, DIARRHEA BLOODY, DIARRHEA CHILLS, DIARRHEA FATIGUE, DIARRHEA HEADACHE, DIARRHEA NAUSEA, DIARRHEA RUNNY NOSE, DIARRHEA VOMITING, DIARRHEA WATERY, DIARRHEA., DIARRHEA. 2021, DIARRHOEA, DIARROHEA, DIFF BREATHING, DIFF SWALLOWING, DIFFICULT BREATHING, DIFFICULT SWALLOWING, DIFFICULTY, DIFFICULTY BREATHING, DIFFICULTY IN BREATHING, DIFFICULTY SWALLOWING, DIGESTIVE ISSUES, DIRECT CONTACT, DIRRHEA, DISCHARGE, DISCOMFORT, DIZINESS, DIZZINES, DIZZINESS, DIZZY, DIZZY HEADACHE, DIZZY NAUSEA, DIZZYNESS, DKA, DOB, DROWSINESS, DROWSY, DRY, DRY COUGH, DRY EYES, DRY MOUTH, DRY NOSE, DRY THROAT, DYSPEPSIA, DYSPHAGIA, DYSPNEA, DYSURIA, Dysuria, EAR, EAR ACHE, EAR ACHES, EAR CONGESTION, EAR INFECTION, EAR PAIN, EAR PRESSURE, EARACHE, EARACHES, EARPAIN, EARS, EARS HURT, EARS PLUGGED, ED, ED PATIENT, ED PT, ELEVATED LIVER ENZYMES OR LIVER FUNCTION TESTS, EMERGE DEPT, EMERGENCY DEPT, EMESIS, EMESIS DIARRHEA, EMESIS X 1, EMESIS X1, EMPLOYEE, EMPLOYEE HEALTH, EMPLOYEE/ STAFF MEMBER, EMS, EMS ASYMP, EMS OUTREACH, EMS SYMP, ENCEPHALITIS, ENDO, EPIGASTRIC PAIN, ER, ER - TO BE HOSPITALIZED, ER ADMIT, ER PATIENT, ER PATIENT ADMITTED, ER PT, ER PT ADMITTED, ERROR, ESSENTIAL CAREGIVER, ESSENTIAL CAREGIVER FOR LONG-TERM CARE, EXHAUSTED, EXHAUSTION, EXPOSED, EXPOSED AND PREGNANT, EXPOSURE, EXPOSURE TO MICE, EXTREME FATIGUE, EXTREME TIREDNESS, EYE, EYE DISCHARGE, EYE INFECTION, EYE IRRITATION, EYE PAIN, EYES, EYES BURNING, EYES HURT, Encephalitis, F, FA, FAILURE TO COPE, FAINT, FAITGUE, FAITUGE, FALL, FALLS, FAT, FATGIUE, FATGUE, FATI, FATIG, FATIGE, FATIGU, FATIGUE, FATIGUE 12, FATIGUE 2021, FATIGUE ACHES, FATIGUE AND CONGESTION, FATIGUE AND HEADACHE, FATIGUE AND NAUSEA, FATIGUE AND RUNNY NOSE, FATIGUE BODY ACHES, FATIGUE CHILLS, FATIGUE CONGESTED, FATIGUE CONGESTION, FATIGUE DIARRHEA, FATIGUE HEADACHE, FATIGUE MALAISE, FATIGUE MUSCLE ACHES, FATIGUE MYALGIA, FATIGUE NASAL CONGESTION, FATIGUE NAUSEA, FATIGUE RUNNY NOSE, FATIGUE SOB, FATIGUE., FATIGUE. 2021, FATIGUE. 2022, FATIGUE. HEADACHE, FATIGUED, FATIGUES, FATIQUE, FATTY LIVER, FATUGUE, FATUIGE, FEB 8 OR, FEBRILE, FEELING FEVERISH, FEELING HOT, FEELING TIRED, FEELING UNWELL, FEELING WARM, FEELING WEAK, FEELS WARM, FELT FEVERISH, FELT WARM, FERTILITY, FEVER, FEVER 37, FEVER 37.8, FEVER 37.9, FEVER 38, FEVER 38.1, FEVER 38.2, FEVER 38.5, FEVER AT HOME, FEVER COUGH, FEVER COUGH SORE THROAT, FEVER RESOLVED, FEVER TODAY, FEVER., FEVER. 2021, FEVERISH, FEVERS, FEVR, FLANK PAIN, FLEM, FLU, FLU LIKE, FLU LIKE SYMPTOMS, FLU SYMPTOMS, FLU-LIKE SYMPTOMS, FLUSHED, FOGGY, FOGGY HEAD, FOR ADMISSION, FOR DISCHARGE, FOR LTC VISIT, FOR OR, FOR PLACEMENT, FOR PRE OP, FOR PRECAUTION, FOR SAFETY, FOR SCHOOL, FOR SURGERY, FOR TRANSFER, FOR WORK, FOR WORK., FULL VAC, FULL VACC, FULL VACC., FULLY VACCINATED., FUSSY, GASTRITIS, GASTRO, GASTRO ISSUES, GASTRO SYMPTOMS, GASTROENTERITIS, GASTROINTESTINAL, GEN UNWELL, GEN WEAKNESS, GENERAL FATIGUE, GENERAL MALAISE, GENERAL UNWELL, GENERAL WEAKNESS, GENERALIZED WEAKNESS, GENERALLY UNWELL, GERD, GI, GI BLEED, GI BLEEDING, GI ISSUE, GI ISSUES, GI SYMPTOMS, GI UPSET, GI bleeding, GOING TO OR, GREEN PHLEGM, GREEN SPUTUM, H, H.VOICE, HA, HA ACHES, HA RN, HA., HADACHE, HALLUCINATIONS, HARD TO BREATH, HARD TO BREATHE, HARD TO SWALLOW, HCV RE-EXPOSURE, HCW, HCW-SURVEILLANCE, HEA, HEAACHE, HEACHACHE, HEACHE, HEAD, HEAD ACHE, HEAD ACHES, HEAD AND BODY ACHE, HEAD COLD, HEAD CONGESTION, HEAD PRESSURE, HEADA, HEADAACHE, HEADAC, HEADACE, HEADACEH, HEADACH, HEADACHE, HEADACHE 9, HEADACHE (ONLY), HEADACHE 12, HEADACHE 2021, HEADACHE ACHES, HEADACHE ACHY, HEADACHE AND BODY ACHE, HEADACHE AND BODY ACHES, HEADACHE AND BODY PAIN, HEADACHE AND CHILLS, HEADACHE AND CONGESTION, HEADACHE AND DIARRHEA, HEADACHE AND FATIGUE, HEADACHE AND NASAL CONGESTION, HEADACHE AND NAUSEA, HEADACHE AND RUNNY NOSE, HEADACHE AND STUFFY NOSE, HEADACHE AND VOMITING, HEADACHE BODY ACHE, HEADACHE BODY ACHES, HEADACHE BODY PAIN, HEADACHE BODYACHE, HEADACHE CHEST PAIN, HEADACHE CHILLS, HEADACHE CONGESTED, HEADACHE CONGESTION, HEADACHE DIARRHEA, HEADACHE DIZZINESS, HEADACHE DIZZY, HEADACHE FATIGUE, HEADACHE FATIGUE NAUSEA, HEADACHE FATIGUED, HEADACHE LOSS OF TASTE, HEADACHE MALAISE, HEADACHE MUSCLE ACHE, HEADACHE MUSCLE ACHES, HEADACHE MUSCLE PAIN, HEADACHE MYALGIA, HEADACHE NASAL CONGESTION, HEADACHE NAUSEA, HEADACHE NAUSEA DIARRHEA, HEADACHE NAUSEA FATIGUE, HEADACHE NAUSEA VOMITING, HEADACHE RHINORRHEA, HEADACHE RUNNY NOSE, HEADACHE RUNNY NOSE FATIGUE, HEADACHE SINUS, HEADACHE SINUS CONGESTION, HEADACHE SNEEZING, HEADACHE SOB, HEADACHE STOMACH ACHE, HEADACHE STUFFY NOSE, HEADACHE TIRED, HEADACHE UPSET STOMACH, HEADACHE VOMITING, HEADACHE VSS, HEADACHE WEAKNESS, HEADACHE., HEADACHE. 2021, HEADACHE. 2022, HEADACHE. CHILLS, HEADACHE. FATIGUE, HEADACHE. NAUSEA, HEADACHE. RUNNY NOSE, HEADACHE.VSS, HEADACHE/STIFF NECK, HEADACHES, HEADACHES BODY ACHES, HEADACHES FATIGUE, HEADACHES RUNNY NOSE, HEADACHES., HEADAHCE, HEADAHE, HEADCAHE, HEADCHE, HEADCOLD, HEALTH CARE WORKER, HEALTHCARE WORKER, HEART FAILURE, HEART PALPITATIONS, HEARTBURN, HEAVINESS IN CHEST, HEAVY BREATHING, HEAVY CHEST, HEAVY HEAD, HEDACHE, HEMATURIA, HEMOPTYSIS, HIA, HIGH RISK, HIGH RISK CONTACT, HIVES, HOARSE, HOARSE THROAT, HOARSE VOI, HOARSE VOICE, HOARSENESS, HOARSENESS OF VOICE, HOMELESS, HORSE VOICE, HOT, HOT AND COLD, HOT AND COLD FLASHES, HOT FLASHES, HR-70, HURTS TO SWALLOW, HYPOXIA, Heart failure, Hematuria, IMMUNOCOMPROMISED, IN LABOUR, IN-PT, INCREASED SOB, INCREASED WOB, INDIGESTION, INDIRECT CONTACT, INFASS, INFASS OUTPATIENT, INFERTILITY, INFORMATION, INPATIENT, INPATIENT (HOSPITALIZED), INPATIENT (ICU, INPATIENT (ICU), INPT, INSOMNIA, INTUBATED, INUIT, IRRITABLE, IRRITATED EYES, IRRITATED THROAT, ITCHY, ITCHY EARS, ITCHY EYES, ITCHY NOSE, ITCHY THROAT, IVF (IN VITRO FERTILIZATION), JAIL, JAN, JAUNDICE, JAW PAIN, JOINT ACHES, JOINT PAIN, JOINT PAINS, KCA, LABOUR, LABOURED BREATHING, LACK OF APPETITE, LACK OF ENERGY, LACK OF TASTE, LACK OF TASTE AND SMELL, LARYNGITIS, LATHARGIC, LBM, LEFT EAR PAIN, LEG PAIN, LETHARGIC, LETHARGY, LETHARY, LIGHT HEADACHE, LIGHT HEADED, LIGHT HEADEDNESS, LIGHT-HEADED, LIGHTHEADED, LIGHTHEADEDNESS, LOA, LONG-TERM CARE VISIT, LOOSE BM, LOOSE BOWEL, LOOSE BOWEL MOVEMENT, LOOSE BOWELS, LOOSE STOOL, LOOSE STOOLS, LOS, LOSE BOWEL MOVEMENT, LOSING VOICE, LOSS, LOSS APPETITE, LOSS OF APETITE, LOSS OF APPETITE, LOSS OF APPITITE, LOSS OF SENSE OF SMELL, LOSS OF SENSE OF TASTE, LOSS OF SENSE OF TASTE AND SMELL, LOSS OF SMELL, LOSS OF SMELL AND TASTE, LOSS OF SMELL OR TASTE, LOSS OF TASTE, LOSS OF TASTE AND SMELL, LOSS OF TASTE AND SMELL., LOSS OF TASTE OR SMELL, LOSS OF TASTE SMELL, LOSS OF VOICE, LOSS SMELL, LOSS SMELL AND TASTE, LOSS TASTE, LOSS TASTE AND SMELL, LOSS VOICE, LOST OF APPETITE, LOST OF SMELL, LOST OF TASTE, LOST OF TASTE AND SMELL, LOST TASTE, LOST VOICE, LOT, LOW APPEPTITE, LOW APPETITE, LOW BACK PAIN, LOW ENERGY, LOW FEVER, LOW GRADE FEVER, LOW RISK, LOWER BACK PAIN, LTC, LTC CLEARANCE, LTC EMPLOYEE, LTC HOME VISIT, LTC PLACEMENT, LTC RESIDENT, LTC SCREEN, LTC SCREENING, LTC VISIT, LTC VISITOR, LTC VISITS, LTC VIST, LTC WORKER, LTCWORKER, LUNG PAIN, LW INFASS, LW INFASS, LW INFASS (ASYMP), LW INFASS (SYMP), LW INFASS ASYMP, LW INFASS OUTPATIENT, LW INFASS SYMP, LW INFASS SYMPT, LW INFASS SYMPT., LW INFASS: COUGH, M, MACULOPAPULAR RASH, MALAISE, MALASIE, MAY 6, MENINGITIS, METALLIC TASTE, MICE DROPPINGS, MIGRAINE, MIGRAINES, MIGRANE, MILD CHEST PAIN, MILD CONGESTION, MILD COUGH, MILD FEVER, MILD HEADACHE, MILD RUNNY NOSE, MILD SOB, MILD SORE THROAT, MUCOUS, MUCUS, MUSCLE, MUSCLE ACHE, MUSCLE ACHES, MUSCLE ACHES FATIGUE, MUSCLE AND JOINT PAIN, MUSCLE FATIGUE, MUSCLE PAIN, MUSCLE PAINS, MUSCLE SORE, MUSCLE SORENESS, MUSCLE WEAKNESS, MUSCLEACHE, MUSCLEACHES, MUSCLES ACHES, MYALGIA, MYALGIA HEADACHE, MYALGIA., MYALGIA. 2021, MYALGIAS, May-06, Meningitis, N, N+V, NA, NARCOTIC ADDICTION, NAS, NAS.CONG, NASAL, NASAL AND CHEST CONGESTION, NASAL CON, NASAL CONG, NASAL CONG., NASAL CONGES, NASAL CONGESITON, NASAL CONGEST, NASAL CONGESTED, NASAL CONGESTI, NASAL CONGESTIO, NASAL CONGESTION, NASAL CONGESTION AND FATIGUE, NASAL CONGESTION AND HEADACHE, NASAL CONGESTION AND RUNNY NOSE, NASAL CONGESTION FATIGUE, NASAL CONGESTION HEADACHE, NASAL CONGESTION RUNNY NOSE, NASAL CONGESTION SNEEZING, NASAL CONGESTION., NASAL CONGESTION. 2021, NASAL CONGESTION. 2022, NASAL CONGESTIONS, NASAL CONGSTION, NASAL CONJESTION, NASAL DISCHARGE, NASAL DRAINAGE, NASAL DRIP, NASAL SYMPTOMS, NASALCONGESTION, NASEAU, NASEL CONGESTION, NASIA, NASUEA, NAU, NAUAEA, NAUS, NAUSA, NAUSE, NAUSEA, NAUSEA AND DIARRHEA, NAUSEA AND FATIGUE, NAUSEA AND HEADACHE, NAUSEA AND VOMITING, NAUSEA AND VOMITTING, NAUSEA CHILLS, NAUSEA DIARRHEA, NAUSEA FATIGUE, NAUSEA HEADACHE, NAUSEA RUNNY NOSE, NAUSEA VOMIT, NAUSEA VOMITING, NAUSEA VOMITING DIARRHEA, NAUSEA VOMITING HEADACHE, NAUSEA VOMITTING, NAUSEA., NAUSEAS, NAUSEATED, NAUSEAU, NAUSEOUS, NAUSIA, NECK PAIN, NEW ADMISSION, NEW ADMIT, NEW SMELL, NG, NIGHT SWEATS, NIL, NO, NO APPETITE, NO COUGH, NO ENERGY, NO FEVER, NO PNEUMONIA, NO SENSE OF SMELL, NO SENSE OF TASTE, NO SMELL, NO SMELL AND TASTE, NO SMELL OR TASTE, NO SOB, NO SORE THROAT, NO SYMPTOMS, NO SYMPTOMS NOTED, NO TASTE, NO TASTE AND SMELL, NO TASTE NO SMELL, NO TASTE OR SMELL, NO VOICE, NON-SPECIFIC SYMPTOM(S) - SURVEILLANCE, NONE, NONE LISTED, NORRHEA, NOSE, NOSE BLEED, NOSE CONGESTION, NOT APPLICABLE, NOT EATING, NOT FEELING WELL, NOT FOR TRAVEL, NOT GIVEN, NOT IMMUNIZED OR INCOMPLETE, NOT PROVIDED, NOT SPECIFIED, NOT VACCINATED, NOV, NS, NSTEMI, NURSING HOME, NURSING STUDENT, NV, NVD, NYD, N\T\V, O, O COVID, O2-97, O2-97%, O2-98, O2-98%, O2-99, O2-99%, OCCASIONAL COUGH, ONSET 20, ONSET 2020, ONSET UNKNOWN, ONSET YYYY-MM-DD, ONSET: 2021, OPP, OPP OFFICER, OR, OR TODAY, OTHER, OTHER (SPECIFY), OUT PT, OUT-PT, OUTBREAK, OUTBREAK INVESTIGATION, OUTPATIENT, OUTPT, OVERDOSE, PAIN, PAIN IN CHEST, PAINS, PALPITATIONS, PANCREATITIS, PARENTS 2X VACC, PART VACC, PATIENT HAVING SURGERY, PEACE OF MIND, PFT, PHELGM, PHLEGM, PHLEGM IN THROAT, PHLEGMY, PHLEM, PINK EYE, PINK EYES, PINKEYE, PLACEMENT, PLUGGED EARS, PND, PNEUMONIA, PNEUMONIA (UNKNOWN), POOR APPETITE, POSITIVE ON RAPID TEST, POSITIVE RAPID, POSITIVE RAPID TEST, POSITIVE RAT, POSSIBLE EXPOSURE, POST NASAL, POST NASAL DRIP, POST PARTUM, PRE CARDIAC CATH, PRE CATH, PRE OP, PRE OP 2020, PRE OP SEPT 16, PRE OP SEPT 30, PRE PROCEDURE, PRE SURGICAL, PRE- OP, PRE-OP, PRE-OP 20, PRE-OP 2021, PRE-PROCEDURE, PRE-SURGICAL, PRECAUTION, PREGNANT, PREGNANT (LABOUR), PREOP, PRESSURE, PRESSURE IN CHEST, PRESSURE IN HEAD, PRESURGICAL, PREVIOUS POSITIVE, PREVIOUS RESULT INDETERMINATE, PROD COUGH, PRODUCTIVE COUGH, PUFFY EYES, Palpitations, R, R NOSE, R.NOSE, RAPID POSITIVE, RAPID TEST POSITIVE, RASH, RASH - NOT SPECIFIED, RASHES, RASPY THROAT, RASPY VOICE, RECEIVED ALL DOSES > 14 DAYS AGO, RECENT FEVER, RECENT TRAVEL, RED EYE, RED EYES, REFUGEE, REGULAR TESTING, REMOTE COMMUNITY, RENAL CLINIC PATIENT, RENAL FAILURE, RENAL FAILURE/RENAL CLINIC PATIENT, REQUIRED BY PUBLIC HEALTH, REQUIRED FOR SURGERY, REQUIRED FOR WORK, RESOLVED, RESPIRATORY SYMPTOMS, RESWAB, RETEST, RETRACTIONS, RETURN TO WORK, RETURNING, RHI, RHINITIS, RHINNORHEA, RHINNORRHEA, RHINO, RHINORHEA, RHINORR, RHINORREA, RHINORRHE, RHINORRHEA, RHINORRHEA CONGESTED, RHINORRHEA HEADACHE, RHINORRHEA-NASAL CONGESTION, RHINORRHEA., RHINORRHEA. 2021, RHIONRHEA, RIGHT EAR PAIN, RINGING IN EARS, RN, RN HA, RNNY NOSE, ROUTINE, RULE OUT COVID 19, RUN NOSE, RUNN YNOSE, RUNNING NOSE, RUNNING NOSE., RUNNING NOSE.VSS, RUNNING NOSR, RUNNNY NOSE, RUNNT NOSE, RUNNU NOSE, RUNNY, RUNNY NOSE, RUNNY AND STUFFY NOSE, RUNNY CONGESTED NOSE, RUNNY EYES, RUNNY N, RUNNY NISE, RUNNY NO, RUNNY NOAE, RUNNY NOE, RUNNY NOISE, RUNNY NOS, RUNNY NOSE, RUNNY NOSE 9, RUNNY NOSE 11, RUNNY NOSE 12, RUNNY NOSE 2021, RUNNY NOSE ACHES, RUNNY NOSE AND BODY ACHE, RUNNY NOSE AND BODY ACHES, RUNNY NOSE AND CHILLS, RUNNY NOSE AND CONGESTED, RUNNY NOSE AND CONGESTION, RUNNY NOSE AND DIARRHEA, RUNNY NOSE AND FATIGUE, RUNNY NOSE AND HEAD ACHE, RUNNY NOSE AND HEADACHE, RUNNY NOSE AND NASAL CONGESTION, RUNNY NOSE AND SNEEZING, RUNNY NOSE BODY ACHE, RUNNY NOSE BODY ACHES, RUNNY NOSE CHEST CONGESTION, RUNNY NOSE CHILLS, RUNNY NOSE CONGESTED, RUNNY NOSE CONGESTION, RUNNY NOSE CONGESTION HEADACHE, RUNNY NOSE COUGH, RUNNY NOSE DIARRHEA, RUNNY NOSE FATIGUE, RUNNY NOSE FATIGUE HEADACHE, RUNNY NOSE HEAD ACHE, RUNNY NOSE HEADACHE, RUNNY NOSE HEADACHE FATIGUE, RUNNY NOSE HEADACHES, RUNNY NOSE HOARSE VOICE, RUNNY NOSE LOSS OF TASTE, RUNNY NOSE MUSCLE ACHES, RUNNY NOSE NASAL CONGESTI, RUNNY NOSE NASAL CONGESTION, RUNNY NOSE NAUSEA, RUNNY NOSE ONSET 20, RUNNY NOSE OR NASAL CONGESTION, RUNNY NOSE OR SNEEZING, RUNNY NOSE SNEEZING, RUNNY NOSE SOB, RUNNY NOSE SORE THROAT, RUNNY NOSE STUFFY NOSE, RUNNY NOSE TIRED, RUNNY NOSE VOMITING, RUNNY NOSE VS NA, RUNNY NOSE WATERY EYES, RUNNY NOSE., RUNNY NOSE. 2021, RUNNY NOSE. 2022, RUNNY NOSE. CONGESTION, RUNNY NOSE. FATIGUE, RUNNY NOSE. HEADACHE, RUNNY NOSE. SNEEZING, RUNNY NOSE. T-36.0, RUNNY NOSES, RUNNY NOSR, RUNNY NOSW, RUNNY NOZE, RUNNY NSOE, RUNNY OSE, RUNNY ROSE, RUNNY STUFFY NOSE, RUNNYNOSE, RUNY NOSE, RUUNY NOSE, RYNNY NOSE, S, SCFHT, SCHIZOPHRENIA, SCHOOL, SCHOOL REQUIREMENT, SCRATCH THROAT, SCRATCHY, SCRATCHY THROAT, SCREENING, SEASONAL ALLERGIES, SEIZURE, SELF ASMT, SELF ASSESSMENT, SELF ASSMT, SEPSIS, SEVERE HEADACHE, SHAKES, SHAKING, SHAKY, SHELTER, SHIVERING, SHIVERS, SHORT BREATH, SHORT OF BREATH, SHORTNESS OF BREATH, SHORTNESS OF BREATH., SHORTNESS OF BREATHE, SHOULDER PAIN, SINUS, SINUS COLD, SINUS CONGESTED, SINUS CONGESTION, SINUS CONGESTION HEADACHE, SINUS HEADACHE, SINUS INFECTION, SINUS ISSUES, SINUS PAIN, SINUS PRESSURE, SINUS SYMPTOMS, SINUSES, SINUSITIS, SLEEPY, SLIGHT COUGH, SLIGHT FEVER, SLIGHT HEADACHE, SLIGHT RUNNY NOSE, SLIGHT SORE THROAT, SLUGGISH, SMELL, SMELL TASTE DISORDER, SNEEZ, SNEEZE, SNEEZING, SNEEZING AND RUNNY NOSE, SNEEZING CONGESTION, SNEEZING HEADACHE, SNEEZING RUNNY NOSE, SNEEZING., SNEEZY, SNEZZING, SNIFFING, SNIFFLE, SNIFFLES, SNIFFLING, SOB, SOB CONGESTION, SOB FATIGUE, SOB HEADACHE, SOB ON EXERTION, SOB RUNNY NOSE, SOB TODAY, SOB UPON EXERTION, SOB., SOBE, SOBOE, SOR ETHROAT, SOR THROAT, SORE, SORE BACK, SORE BODY, SORE CHEST, SORE EAR, SORE EARS, SORE EYE, SORE EYES, SORE JOINTS, SORE LEGS, SORE MUSCLE, SORE MUSCLES, SORE NECK, SORE STOMACH, SORE THOAT, SORE THORAT, SORE THRAOT, SORE THROAT, SORE THROAT 12, SORE THROAT 2021, SORE THROAT ONSET 20, SORE THROAT RUNNY NOSE, SORE THROAT., SORE THROAT. 2021, SORE THROAT. 2022, SORE THT, SORE TUMMY, SORENESS, SORETHROAT, SORETHT, SPUTUM, STAFF, STAFF MEMBER, STEMI, STHROAT, STI, STIFF NECK, STIFFNESS, STOMACH, STOMACH ACHE, STOMACH ACHES, STOMACH BUG, STOMACH CRAMPS, STOMACH DISCOMFORT, STOMACH FLU, STOMACH HURTS, STOMACH ISSUES, STOMACH PAIN, STOMACH PAINS, STOMACH UPSET, STOMACHACHE, STOMACHE, STOMACHE ACHE, STREP THROAT, STROKE, STUFF NOSE, STUFF Y NOSE, STUFFED NOSE, STUFFED UP, STUFFED UP NOSE, STUFFINESS, STUFFING NOSE, STUFFY, STUFFY NOSE, STUFFY AND RUNNY NOSE, STUFFY HEAD, STUFFY NOSE, STUFFY NOSE AND HEADACHE, STUFFY NOSE HEADACHE, STUFFY NOSE RUNNY NOSE, STUFFY NOSE SNEEZING, STUFFY NOSE., STUFFY NOSE. 2021, STUFFY RUNNY NOSE, STUFFYNOSE, SUICIDAL, SURGERY, SURVEILLANCE, SURVIELLANCE, SWAB PRIOR TO CHEMO, SWAB PRIOR TO RADIATION, SWAB PRIOR TO STARTING CHEMO, SWAB PRIOR TO STARTING RADIATION, SWALLOWING, SWEAT, SWEATING, SWEATS, SWEATS AND CHILLS, SWEATY, SWELLING, SWOLLEN EYES, SWOLLEN GLAND, SWOLLEN GLANDS, SWOLLEN LYMPH NODES, SWOLLEN THROAT, SWOLLEN TONSILS, SX, SY, SYM, SYMP, SYMP-COUGH, SYMPOTMATIC, SYMPT, SYMPTOMATIC, SYMPTOMS, SYMPTOMS UNKNOWN, SYNCOPE, Sepsis, Sneezing, Stroke, T, T 36.0, T 36.1, T 36.2, T 36.3, T 36.4, T 36.5, T 36.6, T 36.7, T 36.8, T N, T+0, T+1, T+2, T+2 ENDO, T+3, T+4, T+5, T-35.2, T-35.8, T-36.0, T-36.1, T-36.2, T-36.3, T-36.4, T-36.5, T-36.6, T-N, T35.2, T35.6, T35.8, T35.9, T36, T36.1, T36.2, T36.3, T36.4, T36.5, T36.6, T36.7, T36.8, T36.9, TACHYCARDIA, TACHYPNEA, TASTE, TASTE DISORDER, TEMP, TEMP 36.8, TEMP 36.9, TEMPERATURE, TEMPERATURE 36.8, TEMPERATURE 36.9, TEMPERATURE:, TEMPERATURE: 100, TEMPERATURE: 100.0, TEMPERATURE: 100.1, TEMPERATURE: 100.2, TEMPERATURE: 100.3, TEMPERATURE: 100.4, TEMPERATURE: 100.5, TEMPERATURE: 100.6, TEMPERATURE: 100.7, TEMPERATURE: 100.8, TEMPERATURE: 100.9, TEMPERATURE: 101, TEMPERATURE: 101.0, TEMPERATURE: 101.1, TEMPERATURE: 101.2, TEMPERATURE: 101.3, TEMPERATURE: 101.4, TEMPERATURE: 101.5, TEMPERATURE: 101.6, TEMPERATURE: 101.7, TEMPERATURE: 101.8, TEMPERATURE: 101.9, TEMPERATURE: 102, TEMPERATURE: 102.0, TEMPERATURE: 102.5, TEMPERATURE: 103, TEMPERATURE: 103.0, TEMPERATURE: 104, TEMPERATURE: 35.3, TEMPERATURE: 35.4, TEMPERATURE: 35.5, TEMPERATURE: 35.6, TEMPERATURE: 35.7, TEMPERATURE: 35.8, TEMPERATURE: 35.9, TEMPERATURE: 36, TEMPERATURE: 36.0, TEMPERATURE: 36.1, TEMPERATURE: 36.2, TEMPERATURE: 36.3, TEMPERATURE: 36.4, TEMPERATURE: 36.5, TEMPERATURE: 36.6, TEMPERATURE: 36.7, TEMPERATURE: 36.8, TEMPERATURE: 36.9, TEMPERATURE: 37, TEMPERATURE: 37., TEMPERATURE: 37.0, TEMPERATURE: 37.1, TEMPERATURE: 37.2, TEMPERATURE: 37.3, TEMPERATURE: 37.4, TEMPERATURE: 37.5, TEMPERATURE: 37.6, TEMPERATURE: 37.7, TEMPERATURE: 37.8, TEMPERATURE: 37.9, TEMPERATURE: 38, TEMPERATURE: 38., TEMPERATURE: 38.0, TEMPERATURE: 38.1, TEMPERATURE: 38.2, TEMPERATURE: 38.3, TEMPERATURE: 38.4, TEMPERATURE: 38.5, TEMPERATURE: 38.6, TEMPERATURE: 38.7, TEMPERATURE: 38.8, TEMPERATURE: 38.9, TEMPERATURE: 39, TEMPERATURE: 39.0, TEMPERATURE: 39.1, TEMPERATURE: 39.2, TEMPERATURE: 39.3, TEMPERATURE: 39.4, TEMPERATURE: 39.5, TEMPERATURE: 39.6, TEMPERATURE: 39.7, TEMPERATURE: 39.8, TEMPERATURE: 39.9, TEMPERATURE: 40, TEMPERATURE: 40.0, TEMPERATURE: 40.1, TEMPERATURE: 98, TEMPERATURE: 99, TEMPERATURE: 99.0, TEMPERATURE: 99.1, TEMPERATURE: 99.2, TEMPERATURE: 99.4, TEMPERATURE: 99.5, TEMPERATURE: 99.6, TEMPERATURE: 99.7, TEMPERATURE: 99.8, TEMPERATURE: 99.9, TESTED POSITIVE ON RAPID TEST, THROAT, THROAT CONGESTION, THROAT IRRITATION, THROAT PAIN, THROAT TICKLE, THROWING UP, TICKLE IN THROAT, TICKLE THROAT, TIGHT CHEST, TIGHTNESS, TIGHTNESS IN CHEST, TIGHTNESS IN THE CHEST, TIGHTNESS OF CHEST, TIRED, TIRED HEADACHE, TIRED., TIREDNESS, TO BE ADMITTED, TODAY, TRANSFER, TRANSFER TO LTC, TRAUMA, TRAVEL, TRIEDNESS, TROUBLE BREATHING, TROUBLE SWALLOWING, TUMMY ACHE, U, UK, UN, UN VACC, UNEXP FATIGUE, UNEXPLAINED FATIGUE, UNHOUSED, UNIMMUNIZED OR INCOMPLETE, UNK, UNKNOWN, UNKNOWN COUGH, UNKNOWN FEVER, UNKNOWN PNEUMONIA, UNKNOWN SOB, UNKNOWN SORE THROAT, UNPROTECTED SEX, UNWELL, UPCOMING SX, UPSET STOMACH, UPSET STOMACH HEADACHE, UPSET STOMACHE, URINARY TRACT INFECTION, UTI, V, VACCINATED, VAGINAL BLEEDING, VAGINAL DISCHARGE, VAXXED, VERTIGO, VERY TIRED, VESICULAR RASH, VISIT LTC, VISITING LTC, VITALS STABLE., VOICE, VOICE CHANGE, VOICE LOSS, VOLUNTEER, VOM, VOMIT, VOMIT DIARRHEA, VOMIT X1, VOMITED, VOMITED X 1, VOMITED X1, VOMITIN, VOMITING, VOMITING AND DIARRHEA, VOMITING AND HEADACHE, VOMITING DIARRHEA, VOMITING HEADACHE, VOMITING NAUSEA, VOMITING OR DECREASED DRINKING, VOMITING RUNNY NOSE, VOMITING X1, VOMITING., VOMITING. DIARRHEA, VOMITINGS, VOMITNG, VOMITTED, VOMITTING, VOMITTING DIARRHEA, VOMITTING., VOMMIT, VOMMITING, VOMMITTING, VOMTING, VS NA, VSA, VSS, Vaginal bleeding, WARM, WARM TO TOUCH, WATERY EYES, WEAK, WEAKNESS, WEIGHT LOSS, WET COUGH, WHEEZE, WHEEZING, WHEEZY, WORK, WORK CLEARANCE, WORK REQUIREMENT, WORKS IN LTC, Y, YELLOW PHLEGM, YES, YES COUGH, YES FEVER, YES PNEUMONIA, YES SOB, YES SORE THROAT, YES- NOT SPECIFIED, YM

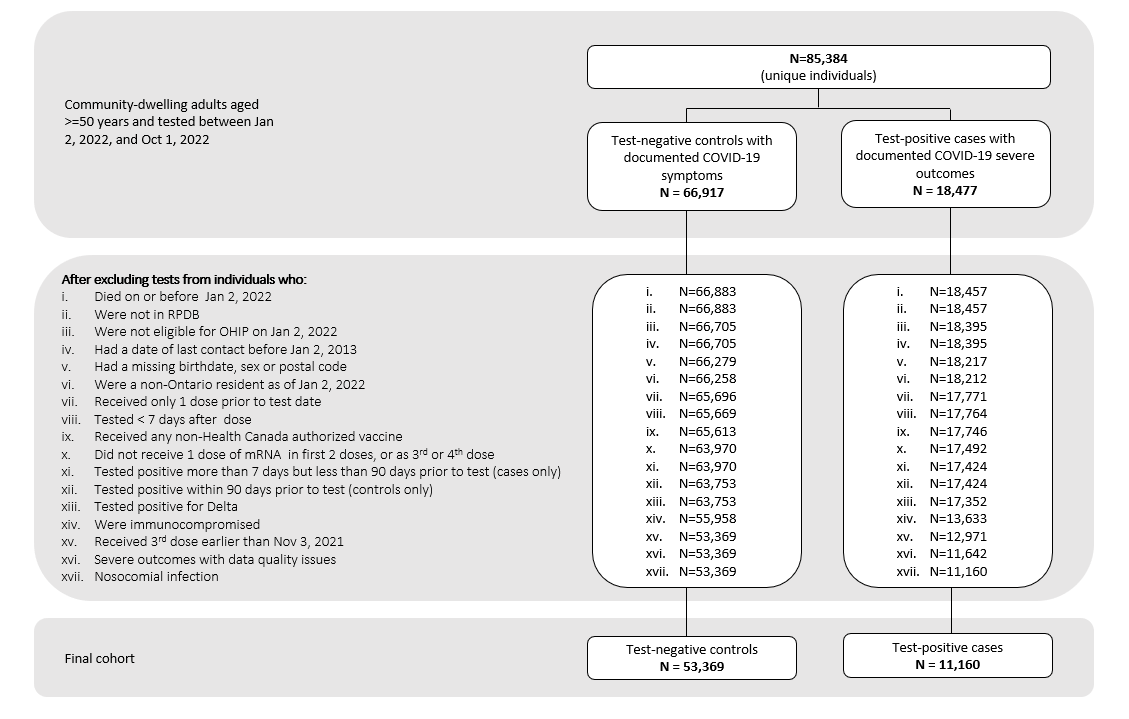

Figure S1: Flow chart of exclusion criteria.

Table S1: List of exposure and covariates used in the analyses

| **Variable** | **Definition** |
| --- | --- |
| Receipt of COVID-19 vaccine | Vaccine information was obtained from COVAXON, the central provincial database compiled and managed by the Ministry of Health. COVAXON contains information on COVID-19 vaccination events for all vaccinations administered in Ontario, including: date(s) of dose administration, reason for administration (e.g., health care worker, long-term care resident, other priority groups), vaccine product information (i.e., manufacturer, lot number, diluent), location and responsible Public Health Unit for vaccination event, and client information. |
| Age | Age was determined from the Registered Persons Database. This variable was included *a priori* as hypothesized to be directly related to COVID-19 infection risk. |
| Sex | Sex was determined from the Registered Persons Database. This variable was included *a priori* as hypothesized to be directly related to COVID-19 infection risk. |
| Public Health Unit region | Taken from Public Health Unit (PHU) information using postal code of residence as recorded in the Registered Persons Database and Statistics Canada Postal Code Conversion File Plus (version 7B). Regions were defined as follows:  Central East: PHU 35 (Haliburton, Kawartha, Pine Ridge District Health Unit), 55 (Peterborough County—City Health Unit), 60 (Simcoe Muskoka District Health Unit)  Central West: PHU 27 (Brant County Health Unit), 34 (Haldimand-Norfolk Health Unit), 36 (Halton Regional Health Unit), 37 (City of Hamilton Health Unit), 46 (Niagara Regional Area Health Unit), 65 (Waterloo Health Unit), 66 (Wellington-Dufferin-Guelph Health Unit)  Durham: PHU 30 (Durham Regional Health Unit)  Eastern: PHU 38 (Hastings and Prince Edward Counties Health Unit), 41 (Kingston, Frontenac and Lennox and Addington Health Unit), 43 (Leeds, Grenville and Lanark District Health Unit), 57 (Renfrew County and District Health Unit), 58 (The Eastern Ontario Health Unit)  North: PHU 26 (The District of Algoma Health Unit), 47 (North Bay Parry Sound District Health Unit), 49 (Northwestern Health Unit), 56 (Porcupine Health Unit), 61 (Sudbury and District Health Unit), 62 (Thunder Bay District Health Unit), 63 (Timiskaming Health Unit)  Ottawa: PHU 51 (City of Ottawa Health Unit)  Peel: PHU 53 (Peel Regional Health Unit)  South West: PHU 31 (Elgin-St. Thomas), 33 (Grey Bruce Health Unit), 39 (Huron County Health Unit), 40 (Chatham-Kent Health Unit), 42 (Lambton Health Unit), 44 (Middlesex-London Health Unit), 52 (Oxford), 54 (Perth District Health Unit), 68 (Windsor-Essex County Health Unit), 75 (Southwestern Health Unit)  Toronto: PHU 95 (City of Toronto Health Unit)  York: PHU 70 (York Regional Health Unit) |
| Weekly period of COVID-19 test | Based on the index date (i.e. specimen collection date, or date of severe outcome if before specimen collection date):   - 2 January to 8 January 2022 - 9 January to 15 January 2022 - 16 January to 22 January 2022 - 23 January to 29 January 2022 - 30 January to 5 February 2022 - 6 February to 12 February 2022 - 13 February to 19 February 2022 - 20 February to 26 February 2022 - 27 February to 5 March 2022 - 6 March to 12 March 2022 - 12 March to 18 March 2022 - 19 March to 26 March 2022 - 27 March 2022 to 2 April 2022 - 3 April to 9 April 2022 - 10 April to 16 April 2022 - 17 April to 23 April 2022 - 24 April to 30 April 2022 - 1 May to 7 May 2022 - 8 May to 14 May 2022 - 15 May to 21 May 2022 - 22 May to 28 May 2022 - 29 May to 4 June 2022 - 5 June to 11 June 2022 - 12 June to 18 June 2022 - 19 June to 25 June 2022 - 26 June to 2 July 2022 - 3 July to 9 July 2022 - 10 July to 16 July 2022 - 17 July to 23 July 2022 - 24 July to 30 July 2022 - 31 July to 6 August 2022 - 7 August to 13 August 2022 - 14 August to 20 August 2022 - 21 August to 27 August 2022 - 28 August to 3 September 2022 - 4 September to 10 September 2022 - 11 September to 17 September 2022 - 18 September to 24 September 2022 - 25 September to 1 October 2022 |
| Prior positive SARS-CoV-2 flag | Laboratory confirmed SARS-CoV-2 by reverse transcription polymerase chain reaction (RT-PCR) >90 days prior to index test records in the Ontario Laboratories Information System (OLIS) |
| Household income quintile | Calculated at the dissemination area (DA) level using Census data by multiplying the median income (before-tax) by the number of households and dividing by the sum of single-person equivalent to obtain income per single person equivalent. |
| Essential workers quintile | Calculated at the DA level, using Census data. For each DA, we calculated the number of individuals ≥15 years old that were working in one of the following Census-defined work categories: Sales and service occupations; trades, transport and equipment operators and related occupations; natural resources, agriculture, and related production occupations; and occupations in manufacturing and utilities.  DAs across the province were then ranked by these percentages into quintiles. |
| Persons per dwelling quintile | The average number of persons in private households, calculated at the DA level using Census data. DAs across the province were ranked by average number of persons per household into 5 categories (quintiles), such that each group contained approximately one-fifth of the DAs. |
| Self-identified visible minority quintile | Calculated at the DA level, using Census data. An individual was marked as “self-identify as a visible minority” if they reported being one or more of the following (wording from the Census): “South Asian (e.g., East Indian, Pakistani, Sri Lankan, etc.), Chinese, Black, Filipino, Latin American, Arab, Southeast Asian (e.g., Vietnamese, Cambodian, Laotian, Thai, etc.), West Asian (e.g., Iranian, Afghan, etc.), Korean, Japanese, or Other—specify”. DAs across the province were then ranked by these percentages into quintiles. |
| Number of COVID-19 tests within 3 months prior to December 14, 2020 | The number of tests for an individual recorded in the ICES-derived COVID-19 Integrated Testing Database (which combines data from the Ontario Laboratories Information System, distributed testing data from laboratories within he COVID-19 Diagnostic Network, and Public Health Case & Contact Management (CCM) solution) between September 14, 2020, and December 14, 2020. |
| Comorbidities | **Chronic Respiratory Disease**  An ICES-specific asthma and COPD database was used to identify patients with chronic respiratory disease.  **Chronic Heart Disease**  Individuals were defined as having chronic heart disease if they had congestive heart failure (as identified through an ICES-derived database)^1^, ischemic heart disease (ICD-10 codes I20, I25, I21, I22 in the past 5 years or CCI procedure codes 1IJ76, 1IJ50, 1IJ54, 1IJ57GQ or CCP procedure codes 481, 4802, 4803 in the past 20 years)^2^, or atrial fibrillation (ICD-9 codes 427.31, 427.32, ICD-10 code I48, or OHIP dxcode 427)^3^ in the past 5 years.  **Hypertension**  An ICES-specific database was used to identify patients with hypertension.^4^  **Diabetes**  An ICES-specific database was used to identify patients with diabetes.^5^  **Immunocompromised**  Individuals were defined as being immunocompromised if they had HIV(as identified through an ICES-specific HIV database)^6^, solid organ transplants [as identified through an ICES-specific database or ICD-10 codes, CCI procedure codes, and OHIP feecodes (codes available upon request)], if they received an allogenic/ autologous bone marrow transplant (CCP procedure code 53.0, CCI procedure codes 1WY19, 1LZ19HHU7, 1LZ19HHU8, OHIP feecode Z426), sickle-cell disease (ICD-10 D57.0 – D57.2; D57.8 or ICD-9 282.6), other immune system disorders (ICD-9 273.2, 279.0, 279.1, 279.2, 279.3, 279.8, 279.9, 289.8; ICD-10 D80, D81, D82, D83, D84, D89; OHIP dxcode 279), received immunosuppressive therapy (>30 days of oral corticosteroid in 6 months before index or receipt of other immunocompromising drugs in the 6 months before index), or active cancer (any of the following treatments in the past 6 months: cancer surgery (codes available upon request), radiation (if the ICD-10 code listed was Z510 in NACRS), chemotherapy (if the ICD-10 code listed was Z511 or Z512 and any evidence of cancer diagnosis in the Ontario Cancer Registry (OCR) prior to the last treatment date) or cancer diagnosis in OCR in the year prior to the index date).  **Autoimmune disease**  An ICES-specific database was used to identify individuals with rheumatoid arthritis, or inflammatory bowel disease. Individuals were considered to have psoriasis if they had 1 hospitalization (ICD-9: 696.1, 696.8; ICD-10: L40.0, L40.1, L40.2, L40.3, L40.4, L40.8, L40.9) or 3 physician billings (OHIP code 696). Individuals were considered to have psoriatic arthritis if they had 1 hospitalization (ICD-9 code 696.0 or ICD-10 codes L40.5, M07.0, M07.1, M07.2, M07.3, M09.0) or 3 physician billings [OHIP dxcode = 721 (at least one of these billings must be billed by a rheumatologist, where spec=48)]. Individuals were considered to have multiple sclerosis if they had 1 hospitalization (ICD-9 code 340 or ICD-10 code G35) or 5 physician billings over 2 years (OHIP dxcode 340).  **Chronic kidney disease**  Diagnosis in DAD, NACRS or OHIP in the past 5 years (ICD-10 codes E102, E112, E132, E142, I12, I13, N08, N18, N19 or OHIP codes 403, 585)^7^, at least 1 dialysis code in each of the 3 months prior to index, or patients who were on chronic dialysis in the year before index date (at least 2 of any of the following codes in OHIP, DAD, or SDS separated by at least 90 days, but less than 150 days OHIP service codes: R849, G323, G325, G326, G860, G862, G865 G863, G866, G330, G331, G332, G333, G861, G082, G083, G085, G090, G091, G092, G093, G094, G095, G096, G294, G295, G864, H540, H740; CCI procedure codes 5195, 6698; CCP procedure code 1PZ21).^8^  **Advanced liver disease**  Identified as patients having cirrhosis [2 or more physician visits (diagnosis code 571), or 1+ hospital diagnosis of cirrhosis (ICD-9 codes 456.1, 571.2, 571.5; ICD-10 codes I85.9, I98.2, K70.3,K71.7, K74.6)] or decompensated cirrhosis [1+ physician visits with diagnosis code 571 and 1+ hospital diagnosis or 1+ procedure (ICD-9 codes 456.0, 456.2, 572.2, 572.3, 572.4, 782.4, 789.5l; ICD-10 codes I85.0, I86.4, I98.20, I98.3, K721, K729, K76.6, K76.7, R17, R18; CCI procedure codes 1.NA.13.BA-FA, 1.NA.13.BA-X7, 1.NA.13.BA-BD, 1.KQ.76GP-NR, 1.OT.52.HA; CCP procedure codes 1006, 6691; OHIP feecode J057, Z591)].^9^  **Dementia**  An ICES-specific database was used to identify patients with dementia.^10^  **Frailty**  Individuals were identified as having medical conditions associated with frailty based on health care encounters recorded in DAD, SDS, NACRS, and OHIP in the 2-years prior to index using the Johns Hopkins ACGⓇ System Version 10.  **History of stroke or transient ischemic attack**  Individuals were identified as having a history of transient ischemic attack if they had 1+ hospitalizations or ED visits with ICD-9 codes 435, 3623 or ICD-10 codes G450, G451, G452, G453, G458, G459, H340. Individuals were identified as having a history of acute ischemic stroke if they had 1+ hospitalization with a main diagnosis coded with ICD-9 codes 434, 436 or ICD-10 codes I63, I64, H34.1. |
| Receipt of home care services | Individuals were defined as receiving home care services if they met the following criteria:  **Short stay:**  Defined in HCD as SRC_admission in (“91”,”92”)  **Long-stay:**  Defined in HCD as “SRC_admission in (“93”,”94”) or SRC_discharge in (“93”,”94”)”.  **Palliative:**  Defined in HCD as SRC_admission in (“95”). |
| Receipt of influenza vaccine | An OHIP billing with any of the following fee codes from October 1, 2019, to September 30, 2020, or October 1, 2020, to September 30, 2021: G590, G591, G592, Q130, Q590, Q690, Q691; or, an ODB billing with any of the following Drug Identification Numbers from October 1, 2019 up to September 30, 2020: 02420643, 02420783, 02432730, 02473283, or October 1, 2020, up to September 30, 2021: 02420643, 02420783, 02432730, 02445646, 02494248, 09857645, 09857646. |

*^1^ Schultz SE, Rothwell DM, Chen Z, Tu K. Identifying cases of congestive heart failure from administrative data: A validation study using primary care patient records. Chronic Diseases and Injuries in Canada 2013;33.*

*^2^ Tu JV, Chu A, Donovan LR, Ko DT, Booth GL, Tu K, et al. The cardiovascular health in ambulatory care research team (CANHEART): Using big data to measure and improve cardiovascular health and healthcare services. Circ Cardiovasc Qual Outcomes 2015;8:204-12.*

*^3^ Tu K, Nieuwlaat R, Cheng SY, Wing L, Ivers N, Atzema CL, et al. Identifying patients with atrial fibrillation in administrative data. Can J Cardiol 2016;32:1561-5.*

*^4^ Tu K, Campbell N, Chen Z-L, Cauch-Dudek KJ, McAlister FA. Accuracy of administrative databases in identifying patients with hypertension. Open Medicine 2007;1:E18-E26.*

*^5^ Hux JE, Flintoft V, Ivis F, Bica A. Diabetes in ontario: Determination of prevalence and incidence using a validated administrative data algorithm. Diabetes Care 2002;25:512-6.*

*^6^ Antoniou T, Zagorski B, Loutfy MR, Strike C, Glazier RH. Validation of casefinding algorithms derived from administrative data for identifying adults living with human immunodeficiency virus infection. PLoS One 2011;6:e21748.*

*^7^ Fleet JL, Dixon SN, Shariff SZ, Quinn RR, Nash DM, Harel Z, et al. Detecting chronic kidney disease in population-based administrative databases using an algorithm of hospital encounter and physician claim codes. BMC Nephrology 2013;14:1-8.*

*^8^ Quinn RR, Laupacis A, Austin PC, Hux JE, Garg AX, Hemmelgarn BR, et al. Using administrative datasets to study outcomes in dialysis patients: A validation study. Medical Care 2010;48:745-50.*

*^9^ Lapointe-Shaw L, Georgie F, Carlone D, Cerocchi O, Chung H, Dewit Y, et al. Identifying cirrhosis, decompensated cirrhosis and hepatocellular carcinoma in health administrative data: A validation study. PLoS One 2018;13:e0201120.*

*^10^ Jaakkimainen L, Bronskill SE, Tierney MC, Herrmann N, Green D, Young J, et al. Identification of physician-diagnosed Alzheimer’s disease and related dementias in population-based administrative data: A validation study using family physicians’ electronic medical records. Journal of Alzheimer's Disease 2016;54:337-49.*

Table S2: Descriptive characteristics of community-dwelling adults aged ≥50 years tested for SARS-CoV-2 between January 2, 2022 and October 1, 2022 in Ontario, Canada, comparing unvaccinated individuals to vaccinated individuals

|  | **Unvaccinated, n (%)^a^** | **2 doses, n (%)^a^** | **SD^b^** | **3 doses, n (%)^a^** | **SD^b^** | **4 doses, n (%)^a^** | **SD^b^** |
| --- | --- | --- | --- | --- | --- | --- | --- |
| **Total** | 6,021 | 15,220 |  | 44,989 |  | 7,810 |  |
| Characteristics |  |  |  |  |  |  |  |
| Age (years), mean (standard deviation) | 70.88 ± 12.60 | 65.52 ± 12.32 | 0.43 | 65.98 ± 12.21 | 0.39 | 75.82 ± 10.93 | 0.42 |
| 50-59 | 1,360 (22.6%) | 6,279 (41.3%) | 0.41 | 17,941 (39.9%) | 0.38 | 558 (7.1%) | 0.44 |
| 60-69 | 1,535 (25.5%) | 3,882 (25.5%) | 0.00 | 11,018 (24.5%) | 0.02 | 1,630 (20.9%) | 0.11 |
| 70-79 | 1,452 (24.1%) | 2,427 (15.9%) | 0.21 | 8,098 (18.0%) | 0.15 | 2,583 (33.1%) | 0.20 |
| ≥80 | 1,674 (27.8%) | 2,632 (17.3%) | 0.25 | 7,932 (17.6%) | 0.24 | 3,039 (38.9%) | 0.24 |
| Male sex | 3,106 (51.6%) | 6,602 (43.4%) | 0.16 | 16,750 (37.2%) | 0.29 | 3,367 (43.1%) | 0.17 |
| Public health unit region |  |  |  |  |  |  |  |
| Central East | 275 (4.6%) | 915 (6.0%) | 0.06 | 3,428 (7.6%) | 0.13 | 567 (7.3%) | 0.11 |
| Central West | 1,050 (17.4%) | 2,498 (16.4%) | 0.03 | 6,937 (15.4%) | 0.05 | 1,325 (17.0%) | 0.01 |
| Durham | 146 (2.4%) | 696 (4.6%) | 0.12 | 2,152 (4.8%) | 0.13 | 204 (2.6%) | 0.01 |
| Eastern | 281 (4.7%) | 609 (4.0%) | 0.03 | 2,215 (4.9%) | 0.01 | 461 (5.9%) | 0.06 |
| North | 808 (13.4%) | 2,565 (16.9%) | 0.10 | 8,726 (19.4%) | 0.16 | 1,674 (21.4%) | 0.21 |
| Ottawa | 152 (2.5%) | 281 (1.8%) | 0.05 | 1,031 (2.3%) | 0.02 | 289 (3.7%) | 0.07 |
| Peel | 811 (13.5%) | 2,151 (14.1%) | 0.02 | 4,561 (10.1%) | 0.10 | 787 (10.1%) | 0.11 |
| South West | 1,147 (19.0%) | 2,692 (17.7%) | 0.04 | 7,991 (17.8%) | 0.03 | 1,273 (16.3%) | 0.07 |
| Toronto | 1,045 (17.4%) | 1,972 (13.0%) | 0.12 | 5,693 (12.7%) | 0.13 | 887 (11.4%) | 0.17 |
| York | 282 (4.7%) | 784 (5.2%) | 0.02 | 2,120 (4.7%) | 0.00 | 299 (3.8%) | 0.04 |
| Missing | 24 (0.4%) | 57 (0.4%) | 0.00 | 135 (0.3%) | 0.02 | 44 (0.6%) | 0.02 |
| Household income quintile |  |  |  |  |  |  |  |
| 1 (lowest) | 1,809 (30.0%) | 3,901 (25.6%) | 0.10 | 8,899 (19.8%) | 0.24 | 1,516 (19.4%) | 0.25 |
| 2 | 1,373 (22.8%) | 3,337 (21.9%) | 0.02 | 9,032 (20.1%) | 0.07 | 1,616 (20.7%) | 0.05 |
| 3 | 1,052 (17.5%) | 3,025 (19.9%) | 0.06 | 8,796 (19.6%) | 0.05 | 1,389 (17.8%) | 0.01 |
| 4 | 982 (16.3%) | 2,692 (17.7%) | 0.04 | 8,864 (19.7%) | 0.09 | 1,507 (19.3%) | 0.08 |
| 5 (highest) | 780 (13.0%) | 2,217 (14.6%) | 0.05 | 9,275 (20.6%) | 0.21 | 1,740 (22.3%) | 0.25 |
| Missing | 25 (0.4%) | 48 (0.3%) | 0.02 | 123 (0.3%) | 0.02 | 42 (0.5%) | 0.02 |
| Essential workers quintile |  |  |  |  |  |  |  |
| 1 (0%–32.5%) | 706 (11.7%) | 1,766 (11.6%) | 0.00 | 7,205 (16.0%) | 0.12 | 1,577 (20.2%) | 0.23 |
| 2 (32.5%–42.3%) | 1,076 (17.9%) | 2,944 (19.3%) | 0.04 | 10,032 (22.3%) | 0.11 | 1,832 (23.5%) | 0.14 |
| 3 (42.3%–49.8%) | 1,254 (20.8%) | 3,303 (21.7%) | 0.02 | 9,816 (21.8%) | 0.02 | 1,722 (22.0%) | 0.03 |
| 4 (50.0%–57.5%) | 1,386 (23.0%) | 3,405 (22.4%) | 0.02 | 9,256 (20.6%) | 0.06 | 1,355 (17.3%) | 0.14 |
| 5 (57.5%–100%) | 1,548 (25.7%) | 3,683 (24.2%) | 0.03 | 8,361 (18.6%) | 0.17 | 1,262 (16.2%) | 0.24 |
| Missing | 51 (0.8%) | 119 (0.8%) | 0.01 | 319 (0.7%) | 0.02 | 62 (0.8%) | 0.01 |
| Persons per dwelling quintile |  |  |  |  |  |  |  |
| 1 (0–2.1) | 1,591 (26.4%) | 3,523 (23.1%) | 0.08 | 10,078 (22.4%) | 0.09 | 2,291 (29.3%) | 0.06 |
| 2 (2.2–2.4) | 1,332 (22.1%) | 3,255 (21.4%) | 0.02 | 9,881 (22.0%) | 0.00 | 1,677 (21.5%) | 0.02 |
| 3 (2.5–2.6) | 802 (13.3%) | 1,964 (12.9%) | 0.01 | 6,300 (14.0%) | 0.02 | 1,038 (13.3%) | 0.00 |
| 4 (2.7–3.0) | 1,130 (18.8%) | 2,980 (19.6%) | 0.02 | 9,664 (21.5%) | 0.07 | 1,601 (20.5%) | 0.04 |
| 5 (3.1–5.7) | 1,112 (18.5%) | 3,370 (22.1%) | 0.09 | 8,739 (19.4%) | 0.02 | 1,125 (14.4%) | 0.11 |
| Missing | 54 (0.9%) | 128 (0.8%) | 0.01 | 327 (0.7%) | 0.02 | 78 (1.0%) | 0.01 |
| Self-identified visible minority quintile |  |  |  |  |  |  |  |
| 1 (0.0%–2.2%) | 1,250 (20.8%) | 3,204 (21.1%) | 0.01 | 10,660 (23.7%) | 0.07 | 1,850 (23.7%) | 0.07 |
| 2 (2.2%–7.5%) | 1,059 (17.6%) | 2,918 (19.2%) | 0.04 | 10,172 (22.6%) | 0.13 | 1,876 (24.0%) | 0.16 |
| 3 (7.5%–18.7%) | 1,031 (17.1%) | 2,567 (16.9%) | 0.01 | 8,165 (18.1%) | 0.03 | 1,601 (20.5%) | 0.09 |
| 4 (18.7%–43.5%) | 1,161 (19.3%) | 2,720 (17.9%) | 0.04 | 7,550 (16.8%) | 0.07 | 1,290 (16.5%) | 0.07 |
| 5 (43.5%–100%) | 1,469 (24.4%) | 3,692 (24.3%) | 0.00 | 8,124 (18.1%) | 0.16 | 1,131 (14.5%) | 0.25 |
| Missing | 51 (0.8%) | 119 (0.8%) | 0.01 | 318 (0.7%) | 0.02 | 62 (0.8%) | 0.01 |
| Receipt of 2019-2020 and/or 2020-2021  influenza vaccination | 1,098 (18.2%) | 5,718 (37.6%) | 0.44 | 25,471 (56.6%) | 0.86 | 6,217 (79.6%) | 1.56 |
| Prior positive SARS-CoV-2 test | 193 (3.2%) | 880 (5.8%) | 0.12 | 1,946 (4.3%) | 0.06 | 165 (2.1%) | 0.07 |
| Number of SARS-CoV-2 tests within 3  months prior to December 14, 2020 |  |  |  |  |  |  |  |
| 0 | 5,438 (90.3%) | 12,143 (79.8%) | 0.30 | 31,987 (71.1%) | 0.50 | 6,192 (79.3%) | 0.31 |
| 1 | 397 (6.6%) | 1,981 (13.0%) | 0.22 | 6,553 (14.6%) | 0.26 | 954 (12.2%) | 0.19 |
| ≥2 | 186 (3.1%) | 1,096 (7.2%) | 0.19 | 6,449 (14.3%) | 0.41 | 664 (8.5%) | 0.23 |
| Any comorbidity | 4,976 (82.6%) | 11,802 (77.5%) | 0.13 | 33,920 (75.4%) | 0.18 | 6,779 (86.8%) | 0.12 |
| Receipt of home care services |  |  |  |  |  |  |  |
| None | 5,634 (93.6%) | 14,257 (93.7%) | 0.00 | 42,559 (94.6%) | 0.04 | 7,303 (93.5%) | 0.00 |
| Short stay | 154 (2.6%) | 417 (2.7%) | 0.01 | 1,193 (2.7%) | 0.01 | 259 (3.3%) | 0.04 |
| Long stay | 207 (3.4%) | 503 (3.3%) | 0.01 | 1,123 (2.5%) | 0.06 | 238 (3.0%) | 0.02 |
| Palliative | 26 (0.4%) | 43 (0.3%) | 0.03 | 114 (0.3%) | 0.03 | 10 (0.1%) | 0.06 |

^*^Note, not unique by person; individuals may be included more than once.

^a^Proportion reported, unless stated otherwise.

^b^SD=standardized difference. Standardized differences of >0.10 are considered clinically relevant. Comparing vaccinated subjects to unvaccinated subjects.

Table S3: Descriptive characteristics of community-dwelling adults aged ≥50 years tested for SARS-CoV-2 between January 2, 2022 and October 1, 2022 in Ontario, Canada, comparing those who received 2 doses of monovalent mRNA COVID-19 vaccines with those with received 3 or 4 doses

|  | **2 doses, n (%)^a^** | **3 doses, n (%)^a^** | **SD^b^** | **4 doses, n (%)^a^** | **SD^b^** |
| --- | --- | --- | --- | --- | --- |
| **Total** | 15,220 | 44,989 |  | 7,810 |  |
| Characteristics |  |  |  |  |  |
| Age (years), mean (standard deviation) | 65.52 ± 12.32 | 65.98 ± 12.21 | 0.04 | 75.82 ± 10.93 | 0.88 |
| 50-59 | 6,279 (41.3%) | 17,941 (39.9%) | 0.03 | 558 (7.1%) | 0.87 |
| 60-69 | 3,882 (25.5%) | 11,018 (24.5%) | 0.02 | 1,630 (20.9%) | 0.11 |
| 70-79 | 2,427 (15.9%) | 8,098 (18.0%) | 0.05 | 2,583 (33.1%) | 0.41 |
| ≥80 | 2,632 (17.3%) | 7,932 (17.6%) | 0.01 | 3,039 (38.9%) | 0.50 |
| Male sex | 6,602 (43.4%) | 16,750 (37.2%) | 0.13 | 3,367 (43.1%) | 0.01 |
| Public health unit region |  |  |  |  |  |
| Central East | 915 (6.0%) | 3,428 (7.6%) | 0.06 | 567 (7.3%) | 0.05 |
| Central West | 2,498 (16.4%) | 6,937 (15.4%) | 0.03 | 1,325 (17.0%) | 0.01 |
| Durham | 696 (4.6%) | 2,152 (4.8%) | 0.01 | 204 (2.6%) | 0.11 |
| Eastern | 609 (4.0%) | 2,215 (4.9%) | 0.04 | 461 (5.9%) | 0.09 |
| North | 2,565 (16.9%) | 8,726 (19.4%) | 0.07 | 1,674 (21.4%) | 0.12 |
| Ottawa | 281 (1.8%) | 1,031 (2.3%) | 0.03 | 289 (3.7%) | 0.11 |
| Peel | 2,151 (14.1%) | 4,561 (10.1%) | 0.12 | 787 (10.1%) | 0.12 |
| South West | 2,692 (17.7%) | 7,991 (17.8%) | 0.00 | 1,273 (16.3%) | 0.04 |
| Toronto | 1,972 (13.0%) | 5,693 (12.7%) | 0.01 | 887 (11.4%) | 0.05 |
| York | 784 (5.2%) | 2,120 (4.7%) | 0.02 | 299 (3.8%) | 0.06 |
| Missing | 57 (0.4%) | 135 (0.3%) | 0.01 | 44 (0.6%) | 0.03 |
| Household income quintile |  |  |  |  |  |
| 1 (lowest) | 3,901 (25.6%) | 8,899 (19.8%) | 0.14 | 1,516 (19.4%) | 0.15 |
| 2 | 3,337 (21.9%) | 9,032 (20.1%) | 0.05 | 1,616 (20.7%) | 0.03 |
| 3 | 3,025 (19.9%) | 8,796 (19.6%) | 0.01 | 1,389 (17.8%) | 0.05 |
| 4 | 2,692 (17.7%) | 8,864 (19.7%) | 0.05 | 1,507 (19.3%) | 0.04 |
| 5 (highest) | 2,217 (14.6%) | 9,275 (20.6%) | 0.16 | 1,740 (22.3%) | 0.20 |
| Missing | 48 (0.3%) | 123 (0.3%) | 0.01 | 42 (0.5%) | 0.03 |
| Essential workers quintile |  |  |  |  |  |
| 1 (0%–32.5%) | 1,766 (11.6%) | 7,205 (16.0%) | 0.13 | 1,577 (20.2%) | 0.24 |
| 2 (32.5%–42.3%) | 2,944 (19.3%) | 10,032 (22.3%) | 0.07 | 1,832 (23.5%) | 0.10 |
| 3 (42.3%–49.8%) | 3,303 (21.7%) | 9,816 (21.8%) | 0.00 | 1,722 (22.0%) | 0.01 |
| 4 (50.0%–57.5%) | 3,405 (22.4%) | 9,256 (20.6%) | 0.04 | 1,355 (17.3%) | 0.13 |
| 5 (57.5%–100%) | 3,683 (24.2%) | 8,361 (18.6%) | 0.14 | 1,262 (16.2%) | 0.20 |
| Missing | 119 (0.8%) | 319 (0.7%) | 0.01 | 62 (0.8%) | 0.00 |
| Persons per dwelling quintile |  |  |  |  |  |
| 1 (0–2.1) | 3,523 (23.1%) | 10,078 (22.4%) | 0.02 | 2,291 (29.3%) | 0.14 |
| 2 (2.2–2.4) | 3,255 (21.4%) | 9,881 (22.0%) | 0.01 | 1,677 (21.5%) | 0.00 |
| 3 (2.5–2.6) | 1,964 (12.9%) | 6,300 (14.0%) | 0.03 | 1,038 (13.3%) | 0.01 |
| 4 (2.7–3.0) | 2,980 (19.6%) | 9,664 (21.5%) | 0.05 | 1,601 (20.5%) | 0.02 |
| 5 (3.1–5.7) | 3,370 (22.1%) | 8,739 (19.4%) | 0.07 | 1,125 (14.4%) | 0.20 |
| Missing | 128 (0.8%) | 327 (0.7%) | 0.01 | 78 (1.0%) | 0.02 |
| Self-identified visible minority quintile |  |  |  |  |  |
| 1 (0.0%–2.2%) | 3,204 (21.1%) | 10,660 (23.7%) | 0.06 | 1,850 (23.7%) | 0.06 |
| 2 (2.2%–7.5%) | 2,918 (19.2%) | 10,172 (22.6%) | 0.08 | 1,876 (24.0%) | 0.12 |
| 3 (7.5%–18.7%) | 2,567 (16.9%) | 8,165 (18.1%) | 0.03 | 1,601 (20.5%) | 0.09 |
| 4 (18.7%–43.5%) | 2,720 (17.9%) | 7,550 (16.8%) | 0.03 | 1,290 (16.5%) | 0.04 |
| 5 (43.5%–100%) | 3,692 (24.3%) | 8,124 (18.1%) | 0.15 | 1,131 (14.5%) | 0.25 |
| Missing | 119 (0.8%) | 318 (0.7%) | 0.01 | 62 (0.8%) | 0.00 |
| Receipt of 2019-2020 and/or 2020-2021  influenza vaccination | 5,718 (37.6%) | 25,471 (56.6%) | 0.39 | 6,217 (79.6%) | 0.94 |
| Prior positive SARS-CoV-2 test | 880 (5.8%) | 1,946 (4.3%) | 0.07 | 165 (2.1%) | 0.19 |
| Number of SARS-CoV-2 tests within 3  months prior to December 14, 2020 |  |  |  |  |  |
| 0 | 12,143 (79.8%) | 31,987 (71.1%) | 0.20 | 6,192 (79.3%) | 0.01 |
| 1 | 1,981 (13.0%) | 6,553 (14.6%) | 0.04 | 954 (12.2%) | 0.02 |
| ≥2 | 1,096 (7.2%) | 6,449 (14.3%) | 0.23 | 664 (8.5%) | 0.05 |
| Any comorbidity | 11,802 (77.5%) | 33,920 (75.4%) | 0.05 | 6,779 (86.8%) | 0.24 |
| Receipt of home care services |  |  |  |  |  |
| None | 14,257 (93.7%) | 42,559 (94.6%) | 0.04 | 7,303 (93.5%) | 0.01 |
| Short stay | 417 (2.7%) | 1,193 (2.7%) | 0.01 | 259 (3.3%) | 0.03 |
| Long stay | 503 (3.3%) | 1,123 (2.5%) | 0.05 | 238 (3.0%) | 0.01 |
| Palliative | 43 (0.3%) | 114 (0.3%) | 0.01 | 10 (0.1%) | 0.03 |

^*^Note, not unique by person; individuals may be included more than once.

^a^Proportion reported, unless stated otherwise.

^b^SD=standardized difference. Standardized differences of >0.10 are considered clinically relevant. Comparing subjects who received 3 or 4 doses to those who received 2 doses.

Table S4: Vaccine effectiveness of 2, 3, and 4 doses of monovalent mRNA COVID-19 vaccines (compared to unvaccinated subjects) against Omicron-associated severe outcomes among community-dwelling adults aged ≥50 years in Ontario, Canada, January 2, 2022 to October 1, 2022

|  | **Age 50-59 years** | | **Age 60-69 years** | | **Age 70-79 years** | | **Age ≥80 years** | |
| --- | --- | --- | --- | --- | --- | --- | --- | --- |
| **Days since last dose** | **Median days** | **VE^a^** | **Median days** | **VE^a^** | **Median days** | **VE^a^** | **Median days** | **VE^a^** |
| Second dose |  |  |  |  |  |  |  |  |
| 7-59 days | 37 | 79 (60, 89) | 40 | 80 (62, 90) | 39 | 84 (57, 94) | 27 | 84 (61, 93) |
| 60-119 days | 97 | 77 (65, 84) | 94 | 81 (69, 88) | 94 | 86 (78, 92) | 94 | 57 (27, 75) |
| 120-179 days | 163 | 85 (79, 89) | 163 | 79 (73, 84) | 164 | 76 (67, 82) | 166 | 60 (47, 70) |
| 180-239 days | 202 | 86 (82, 89) | 202 | 84 (80, 87) | 205 | 69 (62, 75) | 210 | 46 (34, 56) |
| 240-299 days | 268 | 82 (75, 87) | 268 | 72 (64, 78) | 271 | 75 (67, 80) | 268 | 68 (59, 75) |
| ≥300 days | 357 | 77 (69, 83) | 356 | 70 (62, 77) | 361 | 71 (63, 78) | 364 | 64 (55, 71) |
| Third dose |  |  |  |  |  |  |  |  |
| 0-6 days | 4 | 96 (91, 98) | 4 | 95 (91, 97) | 4 | 88 (81, 92) | 3 | 66 (48, 77) |
| 7-59 days | 32 | 97 (96, 98) | 30 | 98 (97, 98) | 34 | 96 (95, 97) | 36 | 91 (89, 92) |
| 60-119 days | 89 | 95 (93, 96) | 90 | 94 (92, 95) | 91 | 92 (90, 94) | 90 | 86 (84, 89) |
| 120-179 days | 145 | 94 (91, 96) | 143 | 91 (88, 93) | 141 | 89 (86, 91) | 142 | 82 (79, 86) |
| 180-239 days | 206 | 88 (83, 92) | 208 | 82 (77, 87) | 208 | 79 (73, 84) | 209 | 75 (68, 80) |
| ≥240 days | 261 | 87 (81, 92) | 260 | 85 (78, 90) | 259 | 79 (71, 85) | 260 | 76 (68, 82) |
| Fourth dose |  |  |  |  |  |  |  |  |
| 0-6 days | 4 | **^b^** | 4 | 98 (91, 99) | 4 | 93 (86, 97) | 4 | 86 (77, 91) |
| 7-59 days | 29 | 97 (91, 99) | 31 | 94 (91, 96) | 31 | 93 (91, 95) | 31 | 92 (90, 94) |
| 60-119 days | 74 | 93 (72, 98) | 84 | 89 (84, 93) | 88 | 92 (89, 94) | 87 | 90 (87, 92) |
| ≥120 days | 142 | 86 (44, 96) | 140 | 88 (79, 93) | 140 | 89 (84, 92) | 147 | 88 (85, 91) |

^a^VE = Vaccine effectiveness. Includes 95% confidence interval.

**^b^**Not reported due to unstable estimate (95% confidence interval width exceeded 100 percentage points).

Table S5: Numbers of vaccinated cases and controls included in the analyses of vaccine effectiveness (2, 3, and 4 doses compared to unvaccinated subjects) and marginal effectiveness (3 or 4 doses compared to 2 doses) of monovalent mRNA COVID-19 vaccines against Omicron-associated severe outcomes among community-dwelling adults aged ≥50 years in Ontario, Canada, January 2, 2022 to October 1, 2022

|  | **Age 50-59 years** | | **Age 60-69 years** | | **Age 70-79 years** | | **Age ≥80 years** | |
| --- | --- | --- | --- | --- | --- | --- | --- | --- |
| **Days since last dose** | **Cases^a^** | **Controls^b^** | **Cases^a^** | **Controls^b^** | **Cases^a^** | **Controls^b^** | **Cases^a^** | **Controls^b^** |
| Second dose |  |  |  |  |  |  |  |  |
| 7-59 days | 10 | 119 | 10 | 60 | 6 | 25 | 10 | 27 |
| 60-119 days | 37 | 396 | 27 | 149 | 27 | 82 | 34 | 32 |
| 120-179 days | 65 | 1,066 | 107 | 544 | 112 | 205 | 125 | 135 |
| 180-239 days | 136 | 2,246 | 206 | 1,230 | 332 | 538 | 542 | 423 |
| 240-299 days | 55 | 848 | 108 | 524 | 133 | 357 | 180 | 290 |
| ≥300 days | 86 | 1,215 | 139 | 778 | 163 | 447 | 363 | 471 |
| Third dose |  |  |  |  |  |  |  |  |
| 0-6 days | 8 | 402 | 14 | 233 | 30 | 109 | 49 | 65 |
| 7-59 days | 68 | 5,681 | 94 | 3,650 | 182 | 2,101 | 299 | 1,496 |
| 60-119 days | 80 | 4,830 | 145 | 2,892 | 243 | 1,994 | 503 | 1,902 |
| 120-179 days | 50 | 3,583 | 117 | 2,104 | 246 | 1,699 | 509 | 1,400 |
| 180-239 days | 73 | 2,086 | 128 | 1,116 | 226 | 816 | 448 | 685 |
| ≥240 days | 40 | 1,040 | 46 | 479 | 109 | 343 | 252 | 324 |
| Fourth dose |  |  |  |  |  |  |  |  |
| 0-6 days | ≤5 | 32 | ≤5 | 95 | 10 | 103 | 29 | 83 |
| 7-59 days | ≤5 | 357 | 28 | 730 | 85 | 979 | 152 | 905 |
| 60-119 days | ≤5 | 119 | 40 | 511 | 106 | 837 | 255 | 835 |
| ≥120 days | ≤5 | 38 | 20 | 204 | 85 | 378 | 237 | 543 |

^a^Number of vaccinated Omicron-positive cases.

^b^Number of vaccinated SARS-CoV-2-negative controls.

Table S6: Marginal effectiveness of 3 or 4 doses of monovalent mRNA COVID-19 vaccines (compared to 2 doses) against Omicron-associated severe outcomes among community-dwelling adults aged ≥50 years in Ontario, Canada, January 2, 2022 to October 1, 2022

|  | **Age 50-59 years** | | **Age 60-69 years** | | **Age 70-79 years** | | **Age ≥80 years** | |
| --- | --- | --- | --- | --- | --- | --- | --- | --- |
| **Days since last dose** | **Median days** | **ME^a^** | **Median days** | **ME^a^** | **Median days** | **ME^a^** | **Median days** | **ME^a^** |
| Third dose |  |  |  |  |  |  |  |  |
| 0-6 days | 4 | 67 (33, 84) | 4 | 68 (41, 82) | 4 | 46 (18, 65) | 3 | 13 (-30, 42) |
| 7-59 days | 32 | 79 (72, 84) | 30 | 87 (83, 90) | 34 | 83 (79, 86) | 36 | 77 (72, 80) |
| 60-119 days | 89 | 74 (66, 81) | 90 | 75 (69, 80) | 91 | 71 (65, 76) | 90 | 66 (61, 71) |
| 120-179 days | 145 | 76 (66, 83) | 143 | 69 (61, 76) | 141 | 64 (56, 71) | 142 | 59 (52, 66) |
| 180-239 days | 206 | 60 (44, 71) | 208 | 52 (37, 63) | 208 | 46 (32, 57) | 209 | 45 (33, 54) |
| ≥240 days | 261 | 57 (35, 72) | 260 | 62 (45, 75) | 259 | 45 (24, 59) | 260 | 48 (33, 60) |
| Fourth dose |  |  |  |  |  |  |  |  |
| 0-6 days | 4 | -**^b^** | 4 | 92 (69, 98) | 4 | 77 (53, 89) | 4 | 67 (46, 80) |
| 7-59 days | 29 | 89 (69, 96) | 31 | 83 (74, 89) | 31 | 80 (74, 85) | 31 | 82 (78, 86) |
| 60-119 days | 74 | 77 (-1, 95) | 84 | 71 (57, 80) | 88 | 78 (71, 84) | 87 | 77 (72, 82) |
| ≥120 days | 142 | -**^b^** | 140 | 69 (47, 82) | 140 | 69 (57, 78) | 147 | 75 (68, 81) |

^a^ME = marginal effectiveness. Includes 95% confidence interval.

**^b^**Not reported due to unstable estimate (95% confidence interval width exceeded 100 percentage points).

Table S7: Vaccine effectiveness against Omicron-associated severe outcomes among community-dwelling adults aged ≥50 years in Ontario, Canada, comparing those who received ≥2 doses of monovalent mRNA COVID-19 vaccines to those who received none, by time since vaccination and age, during BA.1/BA.2 and BA.4/BA.5-predominant periods

| **Age group (years)** | **Days since**  **last dose** | **BA.1/BA.2-predominant period**  **(January 2, 2022 to July 2, 2022)** | | | | **BA.4/BA.5-predominant period**  **(July 3, 2022 to October 1, 2022)** | | | | **Percentage point difference in VE between periods^c^** | **p-value** |
| --- | --- | --- | --- | --- | --- | --- | --- | --- | --- | --- | --- |
|  |  | **Median days** | **Cases^a^** | **Controls^b^** | **VE^c^** | **Median days** | **Cases^a^** | **Controls^b^** | **VE^c^** |  |  |
| 50-59 | Dose 2 (240-299) | 268 | 53 | 759 | 83 (76, 88) | 274 | ≤5 | 89 | 87 (43, 97) | -4 | 0.723 |
|  | Dose 2 (≥300) | 333 | 31 | 629 | 83 (74, 89) | 393 | 55 | 586 | 56 (30, 72) | 27 | **0.003** |
|  | Dose 3 (120-179) | 144 | 40 | 3,351 | 96 (93, 97) | 168 | 10 | 232 | 77 (50, 89) | 19 | **<0.001** |
|  | Dose 3 (180-239) | 192 | ≤5 | 684 | 98 (93, 99) | 213 | 70 | 1,402 | 75 (60, 84) | 23 | **<0.001** |
|  | Dose 4 (7-59) | 28 | ≤5 | 91 | 97 (77, 100) | 29 | ≤5 | 266 | 95 (82, 98) | 2 | 0.653 |
| 60-69 | Dose 2 (≥300) | 327 | 48 | 402 | 79 (71, 86) | 395 | 91 | 376 | 43 (16, 61) | 36 | **<0.001** |
|  | Dose 3 (60-119) | 90 | 144 | 2,859 | 95 (93, 96) | 97 | ≤5 | 33 | 94 (53, 99) | 1 | 0.901 |
|  | Dose 3 (120-179) | 142 | 104 | 1,958 | 92 (90, 94) | 168 | 13 | 146 | 74 (48, 87) | 18 | **0.001** |
|  | Dose 3 (180-239) | 191 | 10 | 233 | 90 (79, 95) | 212 | 118 | 883 | 69 (55, 78) | 21 | **0.007** |
|  | Dose 4 (7-59) | 27 | 14 | 476 | 96 (93, 98) | 39 | 14 | 254 | 88 (78, 94) | 8 | **0.015** |
|  | Dose 4 (60-119) | 69 | ≤5 | 72 | 95 (77, 99) | 86 | 38 | 439 | 81 (70, 88) | 14 | 0.111 |
| 70-79 | Dose 2 (≥300) | 328 | 62 | 223 | 80 (72, 86) | 403 | 101 | 224 | 48 (25, 64) | 32 | **<0.001** |
|  | Dose 3 (7-59) | 34 | 180 | 2,082 | 96 (96, 97) | 41 | ≤5 | 19 | 86 (37, 97) | 10 | 0.083 |
|  | Dose 3 (60-119) | 91 | 239 | 1,962 | 93 (92, 95) | 103 | ≤5 | 32 | 86 (57, 95) | 7 | 0.187 |
|  | Dose 3 (120-179) | 140 | 213 | 1,606 | 92 (90, 94) | 162 | 33 | 93 | 58 (30, 75) | 34 | **<0.001** |
|  | Dose 3 (180-239) | 189 | 20 | 210 | 91 (85, 95) | 214 | 206 | 606 | 59 (44, 70) | 32 | **<0.001** |
|  | Dose 4 (7-59) | 27 | 43 | 689 | 96 (94, 97) | 39 | 42 | 290 | 85 (77, 90) | 11 | **<0.001** |
|  | Dose 4 (60-119) | 70 | 15 | 144 | 93 (87, 96) | 91 | 91 | 693 | 86 (80, 90) | 7 | 0.062 |
| ≥80 | Dose 2 (≥300) | 328 | 148 | 236 | 72 (62, 79) | 406 | 215 | 235 | 40 (18, 56) | 32 | **<0.001** |
|  | Dose 3 (7-59) | 36 | 294 | 1,482 | 92 (90, 93) | 40 | ≤5 | 14 | 76 (27, 92) | 16 | 0.071 |
|  | Dose 3 (60-119) | 90 | 494 | 1,869 | 88 (85, 90) | 99 | 9 | 33 | 83 (61, 93) | 5 | 0.461 |
|  | Dose 3 (120-179) | 141 | 455 | 1,305 | 85 (82, 88) | 165 | 54 | 95 | 64 (44, 77) | 21 | **<0.001** |
|  | Dose 3 (180-239) | 193 | 62 | 184 | 87 (80, 91) | 215 | 386 | 501 | 52 (36, 64) | 35 | **<0.001** |
|  | Dose 4 (7-59) | 29 | 94 | 678 | 94 (92, 95) | 38 | 58 | 227 | 86 (79, 90) | 8 | **0.001** |
|  | Dose 4 (60-119) | 73 | 35 | 233 | 95 (92, 97) | 92 | 220 | 602 | 80 (73, 85) | 15 | **<0.001** |
|  | Dose 4 (≥120) | 133 | 6 | 22 | 92 (78, 97) | 148 | 231 | 521 | 80 (72, 85) | 12 | 0.084 |

Note: Estimates were not reported if they were unstable for either period. Bolded p-values considered significant at p<0.05.

^a^Number of vaccinated Omicron-positive cases.

^b^Number of vaccinated SARS-CoV-2-negative controls.

^c^VE = Vaccine effectiveness. Includes 95% confidence interval.
